# Social thinning and stress generation in childhood ADHD: a systematic review

**DOI:** 10.1101/2025.10.27.25338856

**Authors:** Enrico Venturini, Stephanie Allan, Jala Rizeq, Paul Cannon, Owen Gribbell, Eamon McCrory, Essi Viding, Rachel McCarney, Greta Dertwinkel, Elena McAndie, Gianluca Serafini, Andrea Aguglia, Andrea Amerio, Ruchika Gajwani, Helen Minnis

## Abstract

**Background:** Attention-Deficit Hyperactivity Disorder (ADHD) is a neurodevelopmental condition whose social challenges can be as impairing as its core symptoms. Social thinning (reduction in the number and quality of relationships) and stress generation (increased likelihood of experiencing interpersonal stressors) are key mechanisms underlying psychiatric vulnerability, and might be central to the social challenges in childhood ADHD. We aim to map the existing longitudinal evidence on the interplay between ADHD, these two social processes, and the subsequent psychopathological risk.

**Methods:** This systematic review was conducted in line with PRISMA guidelines (PROSPERO: CRD42023467876). We searched Embase, Medline, CINAHL, and APA PsycINFO from inception to 23 April 2025 for longitudinal observational studies enrolling participants younger than 18 with a formal ADHD diagnosis. The primary outcomes were quantitative associations between ADHD and social thinning or stress generation. Methodological quality was appraised using the Joanna Briggs Checklist for Cohort Studies.

**Findings:** Eleven studies, including 26,177 participants, met inclusion criteria. The overall methodological quality was high, with risk of bias rated as low-to-minimal for ten of the eleven studies. ADHD was consistently associated with social thinning, demonstrated by lower peer acceptance (OR = 5.1) and indirectly by poorer social competence (OR = 7.3), and to stress generation, showing positive associations with aggression (*β* = 0.28) and involvement in bullying (OR = 3.6). These social challenges were associated with a higher incidence of psychiatric disorders, including conduct disorder and major depression.

**Interpretation:** The extant evidence indicates that childhood ADHD is prospectively associated with persistent social thinning and stress generation, from childhood into early adulthood. These mechanisms likely compound the psychosocial impairment from core ADHD traits, contributing to the heightened vulnerability for psychiatric disorders across the lifespan. Clinical interventions for ADHD should adopt a holistic approach that targets social functioning to mitigate long-term adverse outcomes.

**Funding:** None

## Introduction

Attention-Deficit Hyperactivity Disorder (ADHD) is a form of neurodivergence characterised by persistent patterns of inattention and/or hyperactivity–impulsivity, which can significantly impair functioning and development.^1^ Beyond these hallmark traits, ADHD is increasingly recognised for its broader impact on emotional regulation, executive functioning, and social relationships.^2^ Social challenges related to ADHD can be as detrimental to physical and mental well-being as the core traits.^3^

It is already well known that childhood adversity can have a stress-sensitising effect on social functioning:^4,5^ McCrory, Foulkes and Viding presented a compelling transactional model,^6^ in which *social thinning* and *stress generation* are key social mechanisms underlying psychiatric vulnerability in those exposed to adversities such as abuse and neglect.^4^ *Social thinning* refers to an attenuation in the number and quality of relationships over time, while a process in which individuals are more likely to experience interpersonal stressors is called s*tress generation*.^6^

Our hypothesis is that *social thinning* and *stress generation* are more likely to occur in the context of ADHD. For example, traits such as impulsivity and difficulties in sustaining attention might make interpersonal stressor events more likely (*stress generation*)^7,8^ and contribute to difficulties in cultivating and maintaining social connections (*social thinning*).^3,9^ In ADHD, disinhibited and conflict-prone behaviours might, for example, create or maintain interpersonal stress which,^5^ in turn, could lead to an increased likelihood of progressive isolation.^6^ In addition, ADHD’s characteristic difficulties in emotional regulation and social adaptation may perpetuate cycles of persistent stress.^2^ For example, recent longitudinal evidence found strong evidence for the effect of ADHD traits on later social isolation and bullying victimisation.^3,10^

Although several authors have suggested that child abuse and neglect might lead to ADHD, the weight of evidence suggests this is only the case in extreme situations such as severe malnutrition or institutional deprivation.^11^ However, even in less extreme situations, maltreatment is associated with an increase in ADHD traits.^12^ Recent evidence has shown that a high genetic (polygenic) risk for ADHD is associated with an increased risk of experiencing child abuse.^13^ Gajwani and Minnis argued, in their Double Jeopardy model,^14^ that *both* adversity *and* neurodivergence might have a clinically important additive role in increasing the risk of psychiatric disorder (Figure 1), and there is some evidence for this.^15^

**Figure 1.**
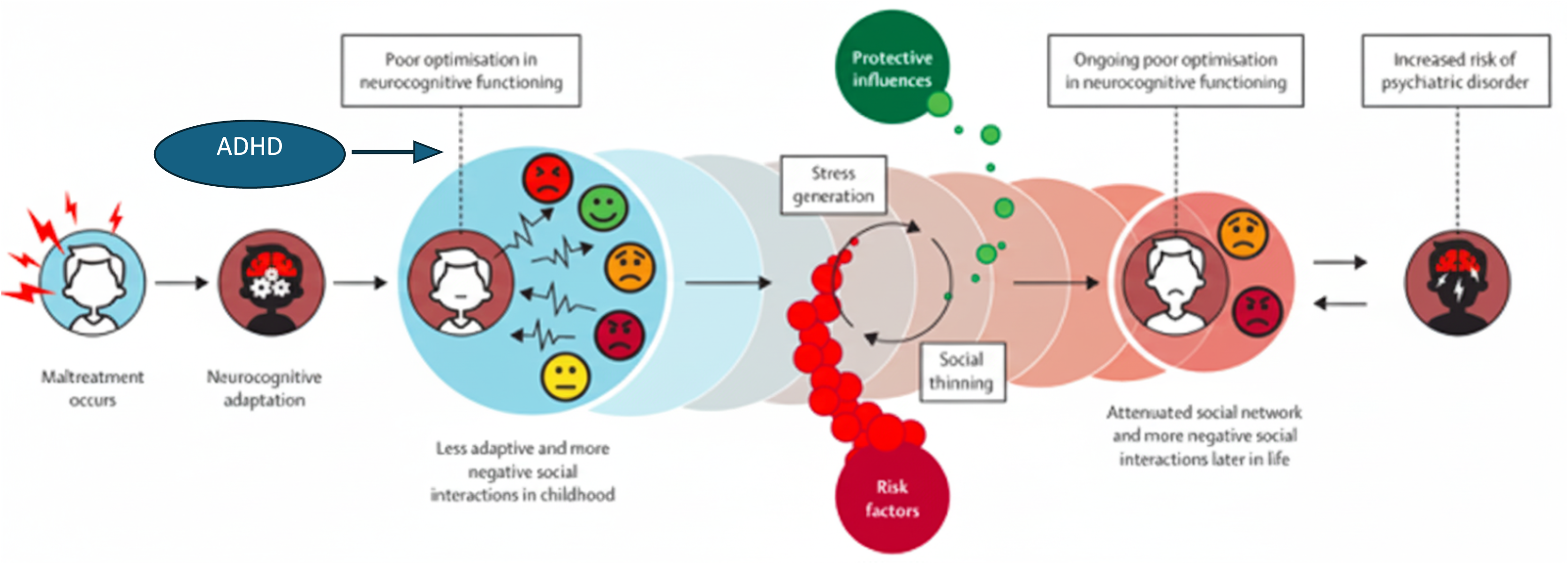
Proposed social transactional model from ADHD to psychiatric disorders, adapted with permission from McCrory et al. (2022). ADHD, by the means of poor optimisation in neurocognitive functioning, could negatively affect social interactions during childhood, prompting stress generation and social thinning, two important social mechanisms perpetuating maladaptive neurocognitive brain functioning and ultimately potentially leading to later psychopathology.

### Rationale and aim of the study

Individuals with ADHD are more likely to experience interpersonal challenges and environmental stressors that intensify maladaptive behaviours, amplifying risks for psychosocial difficulties.^7^ An unfavourable social environment is a well-established correlate of psychiatric vulnerability in people who have experienced childhood trauma.^4–6^ Neurodivergence (including ADHD) is also more common in children who have experienced maltreatment,^3^ and those experiencing traumatic events who are also neurodivergent are at higher risk of developing symptoms of severe mental illness in adolescence.^14^ By mapping the existing evidence, we seek to understand the interplay between ADHD, the two key social processes (*social thinning* and *stress generation*) described above, and the subsequent amplified risk of severe mental illness. Since neurodivergence is increasingly recognised as a crucial factor contributing to psychopathology in people who have experienced childhood trauma,^14^ we believe this addresses an important knowledge gap.

## Methods

### Search strategy and selection criteria

This review is reported in line with the Preferred Reporting Items for Systematic Review and Meta-Analyses (PRISMA) guidelines.^16^ The review protocol (CRD42023467876) was developed by the team and registered on PROSPERO.^17^ Written consent was not required as the analyses involved publicly aggregated data.

The search strategy, designed by PC and SA using a reference set of 25 studies, was reviewed and approved by the team. This search was conducted from inception to 23 April 2025 across the following four databases: Embase, Medline, CINAHL, and APA PsycINFO. It utilised a combination of database-specific subject headings and keywords relating to the diagnosis of ‘ADHD’ and the concepts of ‘social thinning’ and ‘stress generation’. The strategy was adapted for each database to account for differences in subject headings and other functionalities (Appendix 1).

We included longitudinal observational studies published in English enrolling participants younger than 18 years at baseline, with a formal ADHD diagnosis confirmed through clinical assessments or standardised diagnostic instruments. ADHD had to be the primary condition of interest. We excluded animal studies, preprints, literature reviews, trials, qualitative research, as well as investigations focusing solely on ADHD traits or those relying only on parent-reported or non-validated diagnostic measures.

The primary outcomes were the quantitative associations between social thinning and ADHD diagnosis, and between stress generation and ADHD diagnosis, reported as effect sizes. If sufficient data were available, we additionally extracted and reported the frequency of subsequent psychiatric diagnoses and other clinically relevant psychological disturbances as a secondary outcome.

### Data analysis

SA, GD, OG, RMC, EMA and EVe independently screened studies by title and abstract. SA and EVe retrieved full texts for the second screening. Disagreements were resolved by SA, EVe, JR, HM, and RG. EVe, supervised by JR, HM, and RG, extracted data using an ad-hoc spreadsheet without transformation or imputation of the data.

The quality of the papers included was assessed using the Joanna Briggs Checklist for Cohort Studies.^18^ This tool systematically appraises methodological quality across domains such as participant selection, outcome measurement, confounding, and statistical analysis. It provides a criterion-based checklist for determining the RoB through 11 items, each judged as high-risk, low-risk, unclear or not applicable. For this review, item six —‘*Were the groups / participants free of the outcome at the start of the study (or at the moment of exposure)?’*— was deemed not applicable, as the outcomes assessed here vary continuously rather than occurring as discrete events. The remaining ten items were scored and summed to produce a quantitative RoB score, enhancing clarity and reproducibility. This approach was adapted from Glasgow M. J. et al. and is detailed in Appendix 2.^19^ In line with JBI guidance, we also present a qualitative narrative synthesis, recognising that complex criteria cannot be assessed by numerical thresholds alone.^18^

## Results

A total of 14,288 citations were retrieved (Figure 2). Following the removal of duplicates, the titles of 9,161 articles were split among the team (SA, GD, RMC, OG, EVe, JR, RG, HM) for screening for basic exclusion criteria resulting in 252 articles available for full text screening. After removing those without a full-text version, 212 papers were available for full text screening (EVe). Of this, 73 were reviewed by a second (SA) and a third (OG) independent screener. All of the screeners were blind to each other’s work. To ensure screening reliability, Cohen’s kappa was calculated pairwise between the primary screener (EVe) and each independent screener. There was substantial agreement with the second screener (*κ* = 0·75, SE = 0·16, *p* < 0·001) and moderate agreement with the third screener (*κ* = 0·50, SE = 0·13, *p* < 0·001), indicating acceptable inter-rater reliability for study selection decisions. Any disagreement was resolved by consensus discussion. Eventually, eleven studies were available for narrative synthesis.

**Figure 2.**
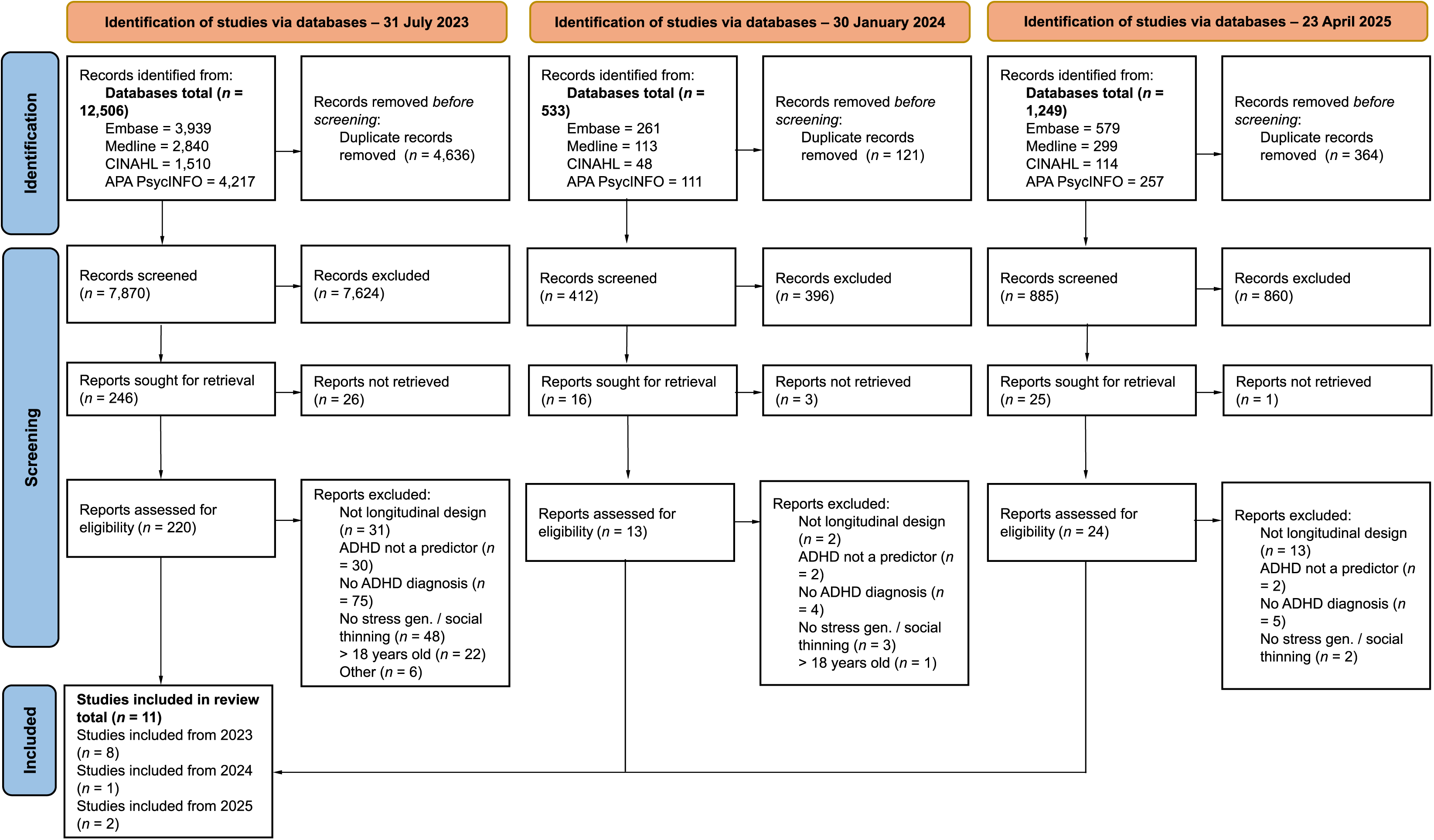
PRISMA 2020 flow diagram for systematic reviews including searches of databases, registers and other sources. The systematic review’s study selection process is illustrated by this flow diagram. It details the identification of records from database searches and other sources, the screening of titles and abstracts, the eligibility assessment of full-text reports, and the eventual inclusion of studies in the review. The number of records at each stage and the reasons for exclusion are also provided.

### Characteristics of studies

The characteristics of the eleven included studies are reported in Table 1. The majority of the studies were conducted in the United States (*n* = 7, 64%). Twenty-six thousand one hundred and seventy-seven participants, from small clinical or community-based samples to large, nationally representative datasets, were followed-up for a minimum of six months up to a maximum of ten years.

**Table 1.** Characteristics of the studies that met inclusion criteria for the systematic review.

| Reference | Country | Baseline sample<br>n (% ADHD) | Baseline age in years<br>range (mean) | Male<br>n (% total) | Follow-up<br>duration | Follow-up sample<br>n (% retained) | ADHD assessment | Outcome | Risk of bias |
| --- | --- | --- | --- | --- | --- | --- | --- | --- | --- |
| Murray-Close D. *et al.*, 2010<sup>37</sup> | US | 820 (65%) | 8–13 (10·4) | 696 (85%) | 6 years | 619 (76%) | Clinical, DISC-IV | ST, SG | ●●○○ |
| Owens E. B. *et al.*, 2009<sup>38</sup> | US | 228 (61%) | 6–12 (9·6) | 0 (0%) | 5 years | 209 (92%) | DISC-IV | ST | ●○○○ |
| Mikami A. Y. *et al.*, 2015<sup>39</sup> | US | 228 (61%) | 6–12 (9·6) | 0 (0%) | 10 years | 217 (95%) | DISC-IV | ST | ●●○○ |
| Lee S. S. *et al.*, 2008<sup>40</sup> | US | 222 (43%) | 4–6 (ns) | 176 (79%) | 8 years | 222 (100%) | DISC | ST | ●●○○ |
| Efron D. *et al.*, 2020<sup>41</sup> | AUS | 477 (56%) | ns (7·3) | 297 (62%) | 3 years | 373 (78%) | DISC-IV | ST | ●●○○ |
| Biederman J., 1996<sup>42</sup> | US | 260 (54%) | 6–17 (11·1) | 260 (100%) | 4 years | 237 (91%) | Clinical, K-SADS-E | ST | ●○○○ |
| Reinke A. L. *et al.*, 2023<sup>43</sup> | US | 231 (52%) | 5–11 (7·9) | 157 (68%) | 6 years | 182 (78%) | DISC-IV | ST | ●○○○ |
| Efron D. *et al.*, 2021<sup>44</sup> | AUS | 372 (47%) | 7–10 (7·3) | 252 (68%) | 18 months | 372 (100%) | DISC-IV | SG | ●○○○ |
| Fogelman N. D. *et al.*, 2018<sup>45</sup> | US | 80 (53%) | 8–12 (9·8) | 47 (59%) | 6 months | 62 (78%) | DISC-IV, VAPRS | SG | ●●○○ |
| Hennig T. *et al.*, 2017<sup>46</sup> | UK | 8,247 (2%) | ns (7·6) | 4,242 (51%) | 5 years | 4,950 (60%) | Clinical, DAWBA | SG | ●●●○ |
| Lin C.-J. *et al.*, 2024<sup>47</sup> | Taiwan | 15,240 (4%) | 13–15 (ns) | 7,948 (52%) | 5 years | 15,240 (100%) | EHR | SG | ●○○○ |

Notes. Risk of bias: ●○○○ = minimal (score 9-10), ●●○○ = low (7-8), ●●●○ = moderate (5-6), ●●●● = high (< 5).
ST = Social Thinning, SG = Stress Generation, K-SADS-E = Schedule for Affective Disorders and Schizophrenia – Epidemiological version, DISC = Diagnostic Structured Interview for Children –IV (DSM-IV criteria) , DAWBA = Development and Well-Being Assessment, VAPRS = Vanderbilt ADHD Parent Rating Scales, EHR = Electronic Health Records, ns = not specified

Overall, gender distribution was almost even, with a slight majority of males (*n* = 14,075, 54%). Two of the included studies (18%) drew data from a sample made up only of girls followed-up prospectively. Ethnicity was inconsistently reported. When specified, samples were predominantly white, with minimal representation of other ethnic groups. In most studies, participants were recruited during childhood, with two studies (18%) including adolescents at baseline. Participants’ ages ranged from 4 to 17 years at baseline and 9 to 24 years at follow-up.

Six out of eleven samples (55%) yielded information on psychiatric disorders, particularly oppositional defiant disorder (ODD), conduct disorder (CD), and internalising disorders (e.g., anxiety, depression). Co-occurring disorders were accounted for as an exclusion criteria or in the analysis, either as covariates or through subgroup analyses. Similarly, information on the use of ADHD medication was variably utilised. Researchers tended to focus more on reporting treatment history, rather than clearly adjusting for medications status.

Risk of bias was low (scores of 7-8) to minimal (9-10) for all except one study, which was of moderate quality (score of 6). Overall, the included studies used robust methodological approaches. Key strengths included adequate control for confounding, and sufficient follow-up periods ranging from six months to ten years. Despite the considerable length of the investigations, in all but one study more than 75% of the sample were retained for follow-up assessments. Recruitment strategies varied, with three studies (27%) recruiting ADHD and comparison groups from different settings (clinical vs. community samples), potentially introducing selection bias (Appendix 3).

### ADHD

ADHD diagnoses were established clinically or with validated instruments, namely the Diagnostic Structured Interview for Children (DISC) being extensively utilised as the tool of choice to ascertain diagnostic status.^20–23^ A Taiwanese study employed data routinely collected at a national level, with no information available on the specific tools aiding the diagnosis, or on the informant-type, but based on at least three validated records, each coded according to the International Classification of Diseases. Overall, over half of the investigations (*n* = 6, 55%) relied on reports from other informants beside the parents (multi-informant). In seven out of eleven cases (64%), a further specification on ADHD presentation was provided. Of note, the focus was primarily on the combined (*n* = 7, 64%) and inattentive (*n* = 5, 46%) types. The hyperactive–impulsive presentation was considered for the analysis in two investigations only (18%) and was, in some cases, an explicit exclusion criterion.

### Social mechanisms and psychiatric disorders

Six studies (55%) focused primarily on social thinning, four (36%) focused on stress generation only, and one (9%) addressed both concurrently. Outcomes were operationalised using a range of subconstructs (Figure 3), which were measured through standardised instruments.

**Figure 3.**
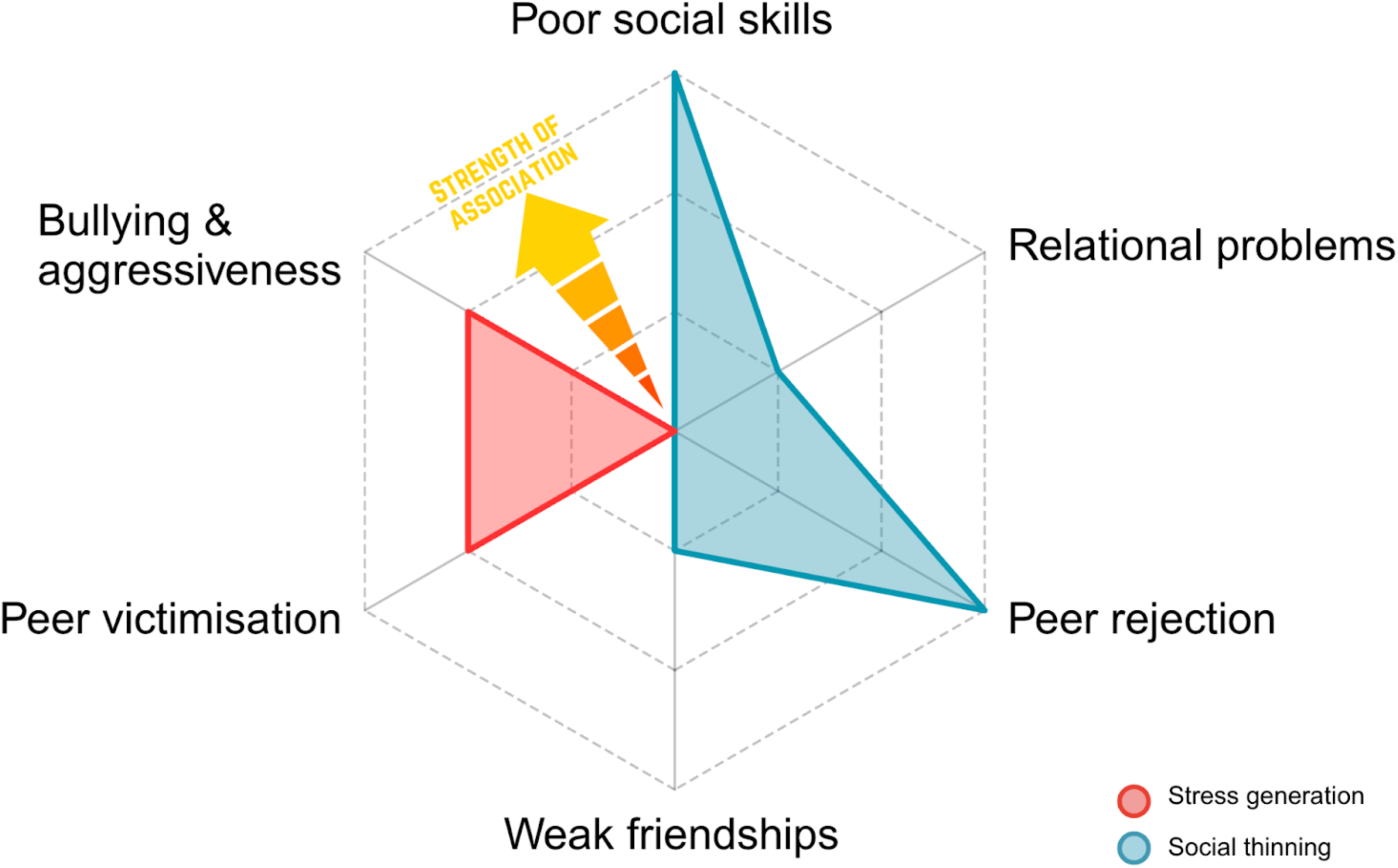
Visual representation of the impairment in social thinning and stress generation in children and adolescents with ADHD. Both social thinning and stress generation encompass several sub-constructs. Each of these sub-constructs is associated with ADHD. The strength of these associations varied: low for relational issues and friendships, moderate for bullying and victimisation, and high for social skills and peer acceptance.

#### Social thinning

Social thinning articulations encompassed interaction skills, relational problems, acceptance from peers and friendship (Table 2). A US study employed trained researchers to directly observe online interplay. Coupled with self-reports and third-party data, this investigation yielded extensive information on the realm of internet-related social problems. The Dishion Social Acceptance Scale was the instrument of choice to measure peer acceptance.^24^ For in-person social competences the Social Skills Rating System (SSRS)^25^ was adopted, whereas for online communication preference —including talking, forming close friendships, or romantic relationships— a 24-item self-report was used.^26^ For relational problems the instruments used were the Social Adjustment Inventory for Children and Adolescents (SAICA),^27^ and the Strength and Difficulties Questionnaire (SDQ)–peer problems subscale.^28^ Friends’ relationships were investigated qualitatively via the Friendship Quality Scale (FQS),^29^ and quantitatively by counting the numbers of friends.

**Table 2.**
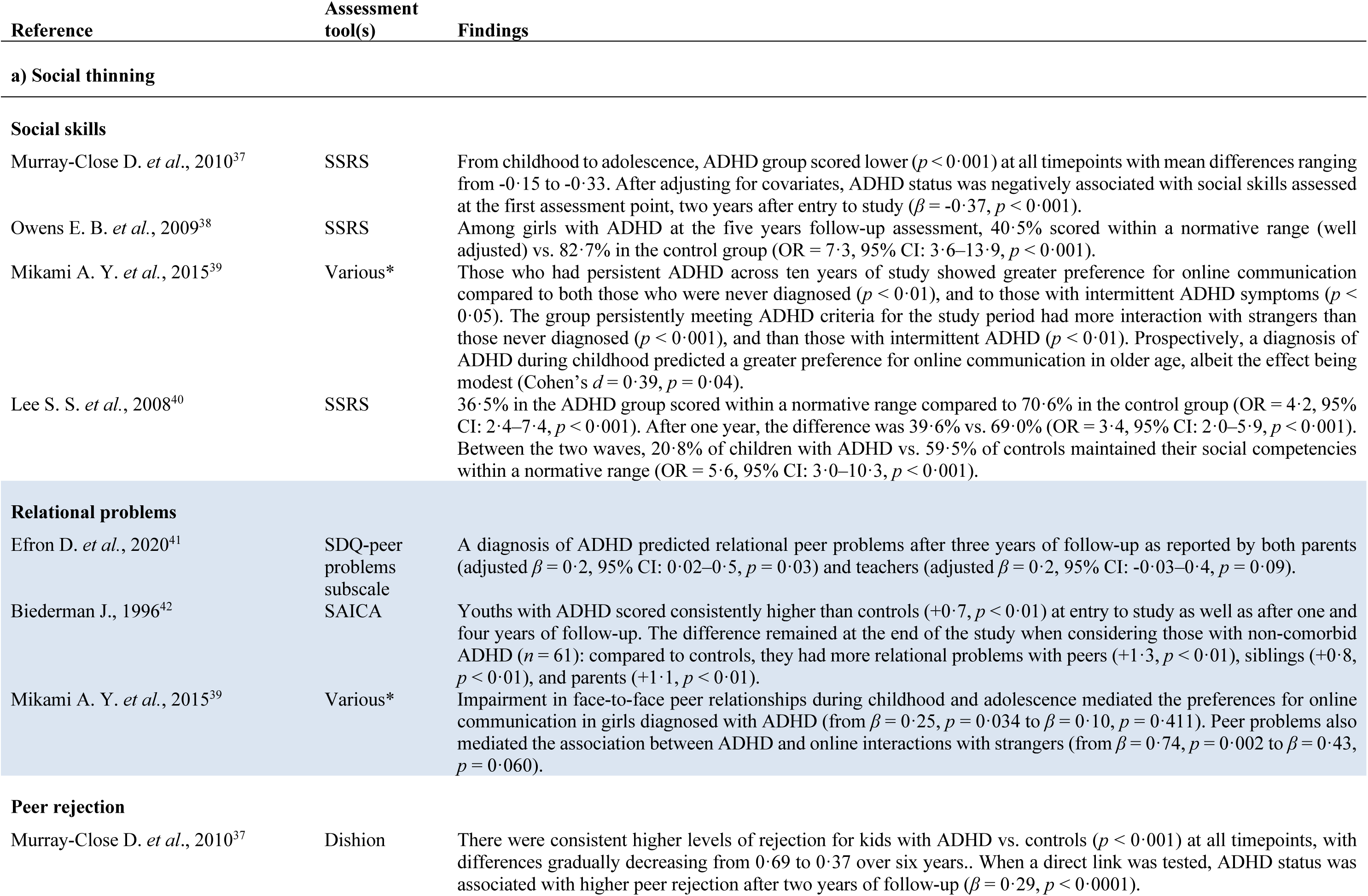
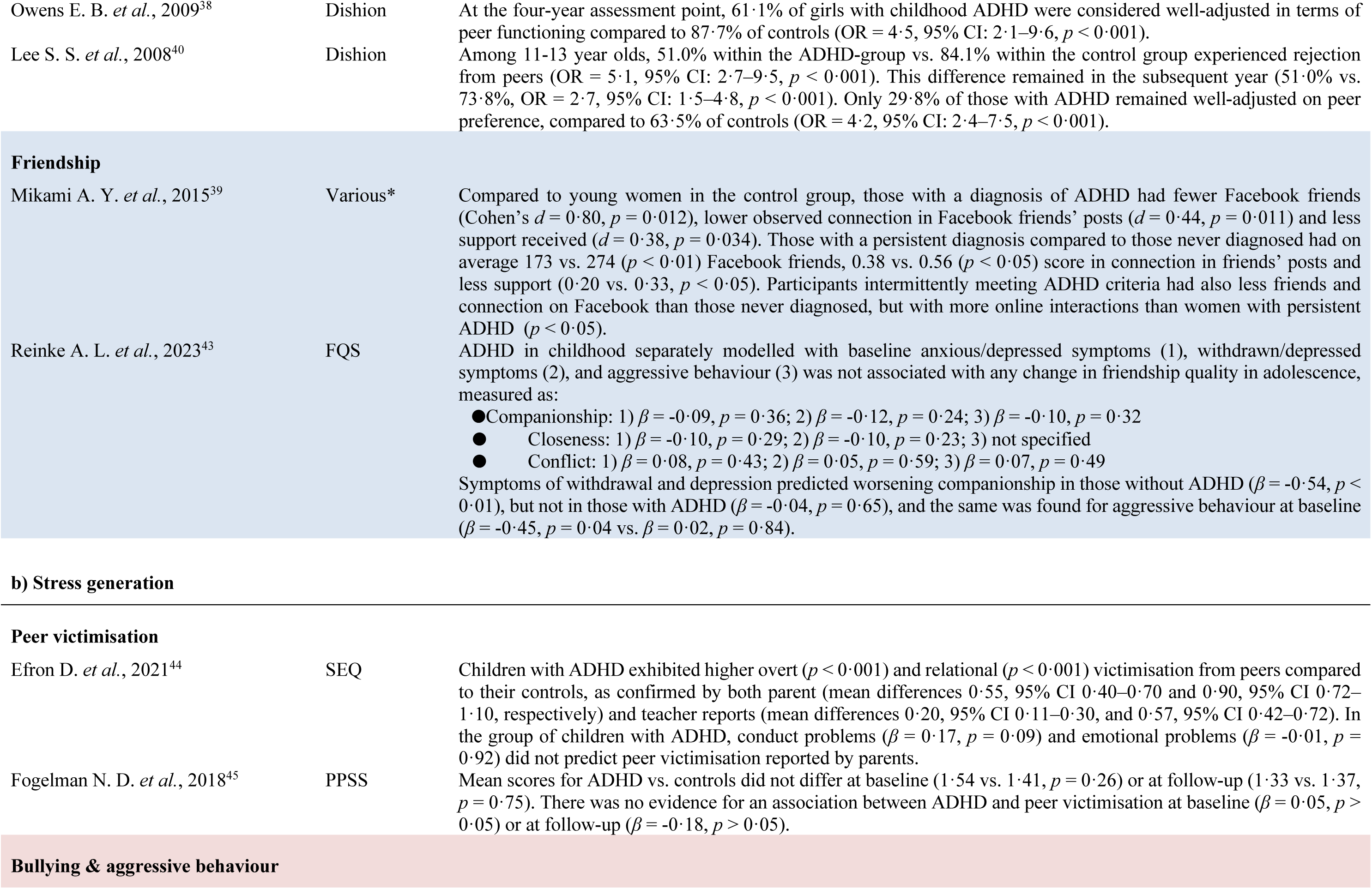
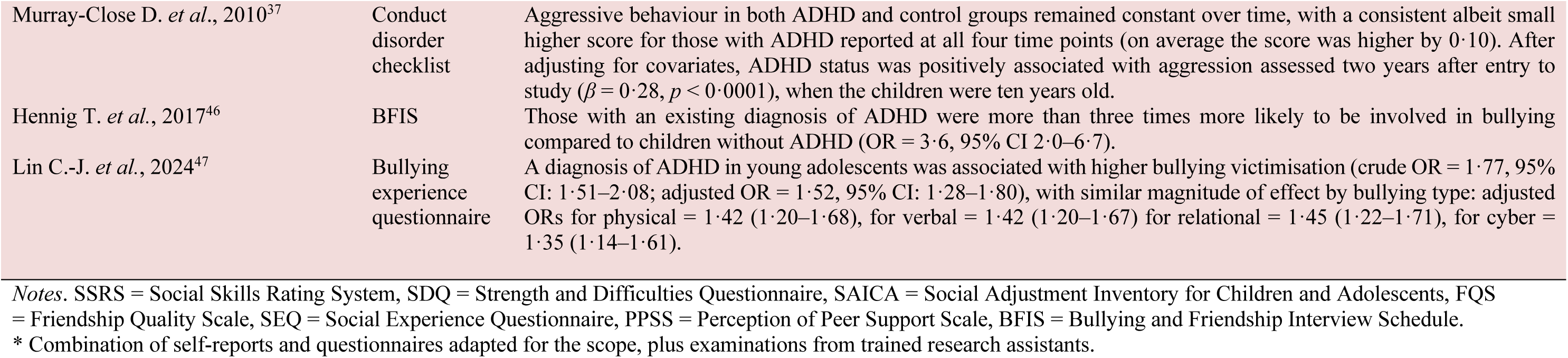
Summary of findings – a) ADHD-social thinning associations, and b) ADHD-stress generation associations.

#### Stress generation

There were two main conceptualisations for stress generation (Table 2). For peer victimisation, both overt and relational, the scales employed were the Social Experience Questionnaire (SEQ)^30^ and the Perception of Peer Support Scale (PPSS).^31^ Aggressiveness was ascertained using the conduct disorder checklist,^32^ and involvement in bullying was assessed via the Bullying and Friendship Interview Schedule (BFIS),^33^ and more in-depth by bullying type using an adapted self-report questionnaire.^34^

#### Psychiatric disorders

Functional consequences and psychiatric disorders emerging in the context of social thinning or stress generation were explored in six studies out of eleven (55%) through the use of several validated scales.^22,23,28,35–37^ The emphasis was mainly on disruptive behaviours and internalising disorders, but bipolar disorder and psychotic and traumatic experiences were also assessed (Table 3).

**Table 3.** ADHD associations with psychiatric disorders.

| Reference | Assessment tool(s) | Findings |
| --- | --- | --- |
| Efron D. *et al.*, 2020<sup>41</sup> | SDQ | ADHD symptom severity at age seven was associated with higher SDQ scores at age ten as reported by parents ($\beta$ adjusted = 0.3, 95% CI: 0.2–0.5, $p = 0.001$) and teachers ($\beta$ adjusted = 0.2, 95% CI: 0.1–0.4, $p < 0.01$). |
| Fogelman N. D. *et al.*, 2018<sup>45</sup> | CBCL | At baseline and follow-up ADHD was correlated with both internalising behaviours ($r = 0.43$, $p < 0.001$ and $r = 0.39$, $p = 0.002$, respectively) and externalising behaviours ($r = 0.62$, $p < 0.001$ and $r = 0.52$, $p < 0.001$). The regression models at baseline and at six months did not show evidence for an association ($\beta = 0.06$, $p = 0.62$ and $\beta = 0.05$, $p = 0.71$, respectively). |
| Lee S. S. *et al.*, 2008<sup>40</sup> | DISC, DBD | At ages 11–13 35.4% of children with preschool ADHD compared to 81.0% of controls were well-adjusted on externalising problems, defined by absence of ODD/CD (OR = 7.7, 95% CI: 4.1–14.3, $p = 0.001$), with similar percentages one year later for the respective groups (33.3% vs. 78.6%, OR = 7.3, 95% CI: 4.0–13.4, $p = 0.001$). Stable well-adjustment across both waves was observed in 23.6% of youths with ADHD vs. 71.4% of controls (OR = 7.7, 95% CI: 4.2–14.0, $p = 0.001$).<br>For internalising problems (anxiety/depression) 67.7% of children with ADHD vs. 90.5% of controls were well-adjusted at the first wave (OR = 4.3, 95% CI: 2.0–9.3, $p = 0.001$), and 63.5% vs. 88.9% were well adjusted at ages 12–14 (OR = 4.6, 95% CI: 2.3–9.2, $p = 0.001$). Stable well-adjustment across waves was observed for 52.8% individuals in the ADHD group vs. 84.1% of those in the control group (OR = 4.7, 95% CI: 2.4–9.1, $p = 0.001$). |
| Mikami A. Y. *et al.*, 2015<sup>39</sup> | DISC-IV | Girls with persistent ADHD showed on average higher levels of self-reported internalising symptoms than consistent comparisons (23.53 vs. 15.77, $p < 0.001$) And also heightened disruptive behaviour symptoms (mean score 9.91 vs. 5.13, $p < 0.001$). |
| Biederman J., 1996<sup>42</sup> | K-SADS-E | 44% of participants with ADHD vs. 7% of controls met criteria for ODD ($p < 0.01$), 14% vs. 0% for CD ($p < 0.01$), 19% vs. 0% for major depression ($p < 0.01$), 9% vs. 0% for bipolar disorder ($p < 0.01$), 17% vs. 1% for social phobia ($p < 0.01$), 11% vs. 1% for separation anxiety ($p < 0.01$), and 18% vs. 0% for tic disorder ($p < 0.01$). |
| Hennig T. *et al.*, 2017<sup>46</sup> | PLIKS, Checklist for traumatic events | Children with ADHD compared to children without ADHD had a more than 3-fold increased risk of psychotic experiences (OR 3.4, 95% CI: 1.8–6.4), and a 2-fold risk of experiencing a traumatic event (OR 2.2, 95% CI: 1.1–4.3). |

*Notes.* SDQ = Strength and Difficulties Questionnaire, CBCL = Child Behavior Checklist, DISC = Diagnostic Interview Schedule for Children -IV (adapted for DSM-IV criteria), DBD = Disruptive Behavior Disorder Rating Scale, K-SADS-E = Schedule for Affective Disorders and Schizophrenia for school-age children Epidemiologic version, PLIKS = Psychosis-Like Symptom Interview, ODD = Oppositional Defiant Disorder, CD = Conduct Disorder.

### Associations between ADHD and subconstructs

#### Social skills

A six-year-long study showed a consistent difference between the ADHD group and controls in terms of social capabilities. From childhood to adolescence, those with ADHD scored lower on the teacher-reported social capabilities at all timepoints. After adjusting for covariates, ADHD status was negatively associated with social skills assessed at time one, two years after entry to study (β = -0·37).^38^ Similarly, a sample of girls with ADHD also demonstrated poorer social competence than comparisons. After five years, among girls with ADHD, 40·5% scored within a normative range (well adjusted) vs. 82·7% in the control group (OR = 7·3, 95% CI: 3·6–13·9).^39^ In early adulthood, those who had persistent ADHD across ten years of study showed greater preference for online communication (over face-to-face) compared to both those who were never diagnosed, and to those with intermittent ADHD symptoms. Moreover, the group who persistently met ADHD criteria throughout the study period had more interactions with strangers —people met online and only known through online communication— than those never diagnosed, or those with intermittent ADHD diagnoses. Prospectively, a diagnosis of ADHD during childhood predicted a greater preference for online communication in older age, albeit the effect being modest (*d* = 0·39).^40^ Challenges in achieving positive-adjustment were consistent at younger ages in a sample composed primarily of male children. At two consecutive assessments one year apart, the odds were up to four times higher in those with ADHD. Moreover, between the two waves, only 20·8% of children with ADHD vs. 59·5% of controls maintained their social competencies within a normative range (OR = 5·6, 95% CI: 3·0–10·3).^41^

#### Relational problems

As reported by both parents and teachers, a diagnosis of ADHD predicted relational peer problems after three years of follow-up (adjusted *β* = 0·2).^42^ This was corroborated cross-sectionally, with those diagnosed with ADHD manifesting more problems with peers than their healthy comparisons. This pattern held true across four years of follow-up and was found also for problems with siblings and parents. The effect remained when comparing individuals with non-comorbid ADHD to controls.^43^ Impairment in face-to-face peer relationships during childhood and adolescence might explain the preferences for online communication in girls diagnosed with ADHD and the higher online interaction with strangers.^40^

#### Peer rejection

The Dishion Social Acceptance Scale was widely employed to investigate differences in peer acceptance. One study spanning six years found consistent higher levels of rejection for kids with ADHD vs. controls.^38^ Four-years forward, among the girls assessed, only 61·1% of those with childhood ADHD were considered well-adjusted in terms of peer functioning, compared to 87·7% of controls (OR = 4·5, 95% CI: 2·1–9·6).^39^ This difference was not limited to females. Among males and females aged 11-13 years, 51·0% within ADHD-group vs. 84·1% within controls (OR = 5·1, 95% CI: 2·7–9·5). Moreover, after one year less than a third of those with ADHD remained well-adjusted on peer preference, compared to almost two thirds of controls.^41^

#### Friendship

Compared to non-ADHD women, analysis of Facebook profiles of young women with ADHD revealed fewer online friends (*d* = 0·80), lower observed connection in Facebook friends’ posts (*d* = 0·44) and less support received (*d* = 0·38).^40^ The findings suggest a gradient effect based on symptom persistence, with the intermittent ADHD group scoring level being in-between the persistent ADHD and comparison group levels. When looking at friendship quality, another study did not find any meaningful association between ADHD diagnostic status in childhood and future adolescent companionship, closeness, or conflict.^44^ Interestingly, ADHD status in childhood seemed to reduce the negative impact of baseline withdrawn/depressed symptoms and aggressive behaviours on friendship quality. Similarly, aggressive behaviour at baseline predicted companionship worsening among those without ADHD but not in the ADHD group.

#### Peer victimisation

Children with ADHD aged seven to nine experienced higher levels of overt (e.g., hitting, pushing, name-calling) and relational (e.g., being excluded, rumours spread about them) victimisation than children without ADHD, as reported by both parents and teachers, even when controlling for conduct and emotional problems.^45^ However, these findings were not replicated in a group of ten-year-olds, albeit the sample was small and the duration of the investigation was six months, that is one third of the previous study.^46^

#### Bullying & aggressive behaviour

In a large sample followed-up for six years the aggressive behaviour in both ADHD and control groups remained constant over time, with a consistent albeit small higher score for those with ADHD reported at all time points. After adjusting for covariates, ADHD status was positively associated with aggression (*β* = 0·28, *p* < 0·0001), assessed when the children were ten years old.^38^ At the same age, another study found that those with an existing diagnosis of ADHD combined subtype were more than three times more likely to be involved in bullying compared to children without ADHD (OR = 3·6, 95% CI: 2·0–6·7).^47^ Nation-wide routine data in Taiwan corroborated the association between ADHD and bullying victimisation (adjusted OR = 1·52, 95% CI: 1·28–1·80), with similar magnitude of effect across bullying types.^48^

### Associations between ADHD and psychiatric disorders

ADHD symptom severity at age seven was associated with higher emotional and behavioural issues at age ten.^42^ Meaningful cross-sectional correlations were observed between ADHD and both internalising and externalising behaviours. However, after controlling for baseline symptom levels, no prospective association with either internalising, or externalising behaviours was found after six months.^46^ Nevertheless, in a much larger investigation, children with preschool ADHD were consistently less well-adjusted in both domains compared to controls.^41^ This difference remained at the early adulthood stage for those with persistent ADHD symptoms, compared to those who consistently remained in the comparison group.^40^ More specifically, in a four-year study, prevalence for several psychiatric comorbidities were higher in those with ADHD compared to controls, particularly for ODD (44% vs. 7%), conduct disorder (14% vs. none), and major depression (19% vs. none).^43^ Moreover, children with ADHD combined subtype compared to children without ADHD had a more than three-fold increased risk of psychotic experiences (OR = 3·4, 95% CI: 1·8–6·4), and a two-fold risk of experiencing a traumatic event (OR = 2.2, 95% CI: 1·1–4·3).^47^

## Discussion

This systematic review provides a comprehensive synthesis of the literature regarding the interplay between ADHD and two key social processes, *social thinning* and *stress generation*, and their association with mental health outcomes. Social thinning and stress generation were evident in the reviewed literature across both peer and family relationships of people with ADHD.

In terms of social thinning, moderate to strong negative associations were found between ADHD and social competence and acceptance from peers,^38–41^ likely arising from ADHD-related behaviours such as intrusiveness, distractibility and failure to respect timing during conversations.^7^ These challenging real-world peer relations might explain a greater preference for virtual communication platforms.^7,40^

Stress generation in peer relations was also found to be associated with ADHD, which was positively associated with aggressive behaviours, both concurrently,^3^ and prospectively.^38^ Negative outward behaviour toward other children, coupled with scarce insight into their behaviours (or positive illusory bias), might likely place these children at heightened risk of experiencing multiple forms significant interpersonal stress including bullying (verbal, physical, relational and cyberbullying).^47,48^ Although children with ADHD can act also as perpetrators (a form of stress generation),^47^ the consensus is that they are more likely to be victims of bullying than their neurotypical peers —likely an example of social thinning arising from stress generation.^8,45–48^

In family relationships, childhood ADHD is associated with relational problems with siblings and parents.^43^ This is consistent with a pattern of stress generation arising from ADHD traits such as impulsivity and poor concentration and the anger and frustration that is associated with a genetic risk for ADHD.^2,9^ Family genetics might also play an important role,^2,49^ since other family members may also have ADHD feature and may be facing similar neurocognitive and psychosocial challenges.^7,13^

Taken together, ADHD is associated with a significant and persistent negative impact on social functioning, in both family and peer relationships, from childhood to adulthood. This could compound the existing psychosocial impairment caused by its core traits, contributing to the heightened vulnerability for psychiatric problems.^6^

A deeper understanding of stress generation and social thinning in ADHD could inform support for changes in children’s environment that could positively affect their functioning and complement treatments targeting core symptoms, which may themselves have positive social consequences.^2,9^ Future investigations should focus on the effect of social processes, adjusting adequately for treatment type, developmental timing of medication, and concurrent neurodivergence or psychiatric disorders. In order to expand the research scope, sub-threshold cases should also be considered, given that individuals who do not persistently meet ADHD diagnostic criteria are also at risk of experiencing social difficulties,^40^ and early identification in these cases may help prevent worsening outcomes.^42^ This aligns with the conceptualisation of ADHD as the extreme expression of a continuous dimension, rather than having clear diagnostic boundaries.^2,49^

Moreover, further research endeavours should test the mediating role of social thinning and stress generation on the subsequent heightened psychiatric risk, which may be further intensified by early exposure to abuse and neglect. Importantly, child maltreatment or mis-attuned care should also be considered, given its association with ADHD “symptom load”,^14^ the independent association between child maltreatment and later aggression and psychopathology.^2^ Current recommendations advocate for a comprehensive assessment including psychosocial and environmental stressors.^1,3^ Considering the complex interplay between ADHD and social factors such as family and peer relationships,^7,9,50^ it will be important to explore whether these factors moderate the path from ADHD to later adverse outcomes. For instance, higher social support has been linked to lower emotional dysregulation in adults with ADHD.^51^

By intervening to reduce the likelihood of social thinning and stress generation there could be a reduction in the sustained stress that can result from the mismatch between individual’s expectations and behaviours and the surroundings.^3,6^ This could translate to better psychosocial functioning and improved control of core ADHD traits breaking the cycle that perpetuates major mental health concerns (Figure 4). Considering the interaction between ADHD, stress and social environment, fostering a stable and supportive relational network proves important, particularly when also childhood adversity is present.^5^

**Figure 4.**
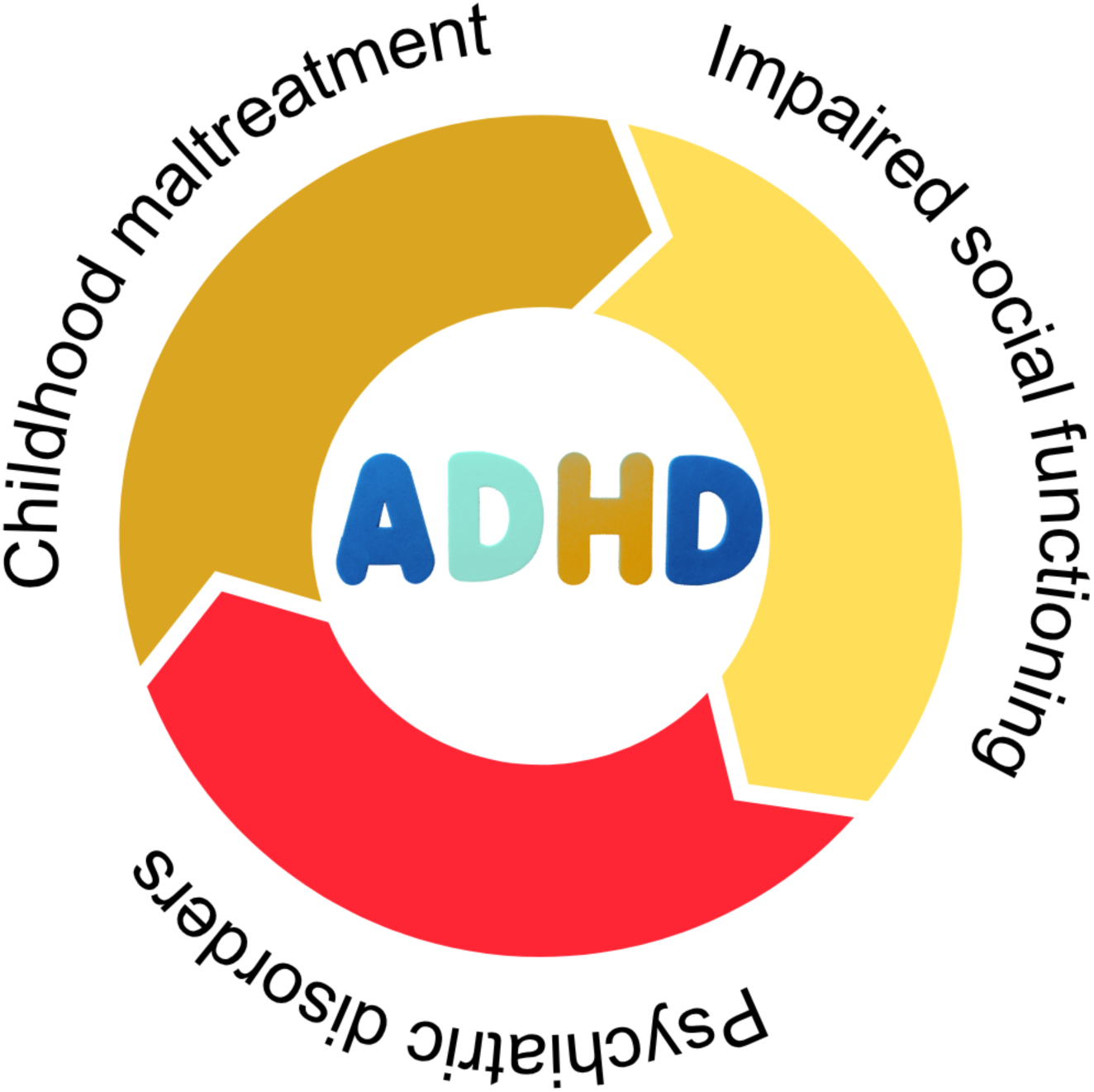
Cyclical relationship between child maltreatment, social impairment, and psychiatric disorders, with ADHD as a key contributing and connecting factor. Early experiences of maltreatment could negatively affect the social sphere, increasing the risk of psychopathology. Both ADHD and childhood maltreatment may lead to cognitive and social dysfunction. The core and auxiliary symptoms of ADHD can further elevate the likelihood of developing psychiatric disorders.

## Strengths and Limitations

Inclusion of longitudinal studies spanning up to ten years duration offers a thorough, long-term view. The use of validated measures for both outcomes and ADHD ascertainment strengthens the credibility of the findings,^20–37^ and the large sample size (2,352 ADHD participants out of 26,177) supports their broad applicability. In addition, in most cases (55%) ADHD status was confirmed through probes collected from more than one informant, in line with best-practice clinical recommendations.^50^

With the exception of two studies, all included participants exhibited either the inattentive or combined presentation of ADHD, when this was specified. While this necessitates caution in generalising the findings, it could also mask larger effects, given that hyperactive–impulsive symptoms tend to be more strongly associated with social isolation.^3^

An additional limitation was the inconsistent inclusion of treatment data in the analyses. Since treatment is usually beneficial for social functioning,^8^ failure to adjust for treatment effects — particularly when most participants are receiving treatment— could lead to underestimates of the true strength of associations with ADHD. Nevertheless, this aspect is understandable in studies with extended follow-up periods, where adherence to care regimens is challenging to gauge.

Importantly, the broader mechanisms of stress generation and social thinning are represented by a variety of distinct but related subconstructs, such as peer acceptance, friendship, and bullying experiences. This complexity makes it challenging to investigate the overarching social mechanisms directly. Combined with considerable methodological heterogeneity across the included studies, this meant a meta-analysis was not feasible. Therefore, effect sizes have been best interpreted within each specific subconstruct.

In conclusion, the extant evidence is consistent with a positive prospective association between ADHD and both social thinning and stress generation. We suggest that this may contribute to the well-known association between ADHD and a higher psychiatric burden across the lifespan.

The literature evidenced consistent social disadvantage experienced by youths diagnosed with ADHD, exemplified by both *social thinning* and *stress generation*, persisting until early adulthood and associated with mental ill-health. This leads us to suggest a simple model that, we hope, will encourage clinical consideration of potential for childhood adversity, social difficulties, and the increased risk of psychiatric disorder in children with ADHD (Figure 4). There are important clinical implications arising from this model:

● Children with ADHD (including those who have not been maltreated) are at higher risk than their peers of experiencing stress generation and social thinning and may need specific support with developing and maintaining more positive social relationships.
● Children with ADHD who have shown patterns of social thinning and stress generation are at higher risk than their peers of experiencing maltreatment or bullying/victimisation and these risks might be ameliorated by targeted focus on their social functioning and emotion regulation.
● Maltreated children may be experiencing stress generation and social thinning arising from *both* trauma-related neurocognitive adaptations *and* from ADHD traits. It is therefore crucial that *both* trauma-related symptomatology (e.g. distorted threat perception, indiscriminate behaviours) and ADHD traits (e.g. impulsivity) are considered as potential intervention targets in maltreated children.

We recommend that clinicians adopt a holistic approach when assessing individuals with ADHD, moving beyond the core features of inattention and hyperactivity–impulsivity. Clinicians should first understand the multifaceted social profile (Figure 3) resulting from the interplay of the various systemic subconstructs, such as the wider relational and familial support networks, to the more individual level factors such as the experiences of peer victimisation. Secondly, once there is a joint shared understanding, it is important to align the individual’s internal scheme of social interactions with the relational context experienced. The focus should be on adapting the relational context by fostering the social capacities of neurodivergent children on the one hand, and by cultivating understanding and support for ADHD among family, friends, and society at large.

## Contributors

Authors SA, HM and RG designed the study and wrote the protocol with help from PC and EVe. Studies were independently reviewed for eligibility by title and abstract (SA, GD, OG, RMC, EMA and EVe). Full-text screening and data extraction was performed by EVe, with supervision from JR, HM, and RG. Authors EVe, SA, JR, EMC, EVi, HM and RG wrote the first draft of the manuscript. The final version of the manuscript was thoroughly revised by AAg, AAm, GS, and then approved by all authors.

## Data sharing

This analysis draws exclusively on data extracted from published studies. No new or individual-level data were collected or generated for this review. The corresponding authors will provide the data extraction forms and extracted data that support this systematic review, upon reasonable request.

## Data Availability

The corresponding authors will provide the data extraction forms and extracted data that support this systematic review, upon reasonable request.

