## Appendix for "Social thinning and stress generation in childhood ADHD: a systematic review"

### Appendix 1

Search strategy

First search, 31 July 2023

**Source: Embase 1947-Present, updated daily**
Interface: OvidSP

Database coverage dates: 1947 to present

Search date: 31 July 2023

Retrieved records: 3,939

Search strategy:

1  attention deficit hyperactivity disorder/ (9280)
2  ((attention* or behav*) adj3 (defic* or disorder* or dysfunc*)).tw. (106434)
3  adhd.tw. (45108)
4  "ad/hd".tw. (500)
5  addh.tw. (149)
6  adhs.tw. (710)
7  or/1-6 (122841)
8  exp child/ (3494474)
9  adolescent/ (1912686)
10  (child* or boy* or girl* or toddler* or infant* or pre?school* or schoolchild* or schoolgirl* or schoolboy* or adolescen* or juvenile* or teen* or young* person* or young* people* or youth* or minor?).tw. (3577598)
11  "minor (person)"/ (939)
12  exp pediatrics/ (139460)
13  (pediatric* or paediatric*).tw. (713328)
14  or/8-13 (5671981)
15  (stress* adj2 (event* or generat* or dependent or minor or major)).tw. (33656)
16  oxidative.tw. (493988)
17  15 not 16 (27728)
18  daily hassle?.tw. (785)
19  social structure/ (6281)
20  social competence/ (7679)
21  social support/ (114357)
22  integration/ (7289)
23  exp social isolation/ (34172)
24  social capital/ (3955)
25  (social adj (thinning or structure or architecture or group? or experience? or skill? or support or ratings or status or integrat* or relationship? or exclusion or contact? or dysfunction* or depriv* or isolat* or capital)).tw. (136177)
26  SNA.tw. (4929)
27  personal network.tw. (256)
28  (social adj2 process*).tw. (9188)
29  socialising.tw. (487)
30  ((interpersonal or affiliative or supportive) adj1 relationship?).tw. (9620)
31  (shared adj (understanding or norms or values or identit*)).tw. (2375)
32  community participation/ (4254)
33  (community adj (participation or inclusion or involve*)).tw. (6808)
34  "Involvement in a life situation".tw. (20)
35  participation.tw. (245138)
36  ((formal or intimate or close) adj relationships).tw. (6491)
37  friendship/ (4333)
38  friend*.tw. (154501)
39  ((rejected or excluded or disliked) adj children).tw. (691)
40  ((peer? or inter?personal) adj2 (accept* or problem? or conflict or likability or liked or status or interact* or function* or relationship? or interaction? or reject*)).tw. (24944)
41  ((number? or quality or quantity) adj2 relationship?).tw. (9232)
42  bullying/ (10355)
43  (victim* or bullied or bully*).tw. (87468)
44  marginali#ed.tw. (10072)
45  loneliness/ (14532)
46  lonel*.tw. (15985)
47  (popular* or unpopular*).tw. (172229)
48  social network/ or social network analysis/ (24401)
49  social network.tw. (14255)
50  online social network/ or social media/ (48099)
51  (48 or 49) not 50 (26861)
52  or/17-47,51 (960101)
53  7 and 14 and 52 (5254)
54  animal/ (2128319)
55  human/ (26565541)
56  54 not (54 and 55) (1614728)
57  53 not 56 (5246)
58  limit 57 to english language (4892)
59  limit 58 to "remove preprint records" (4881)
60  exp "review"/ (3190965)
61  exp clinical trial/ (1844916)
62  exp qualitative research/ (117237)
63  or/60-62 (4954561)
64  59 not 63 (3939)

**Source: Ovid MEDLINE(R) ALL**
Interface OvidSP

Database coverage dates: 1946 to 28 July 2023

Search date: 31 July 2023

Retrieved records: 2,840

Search strategy:

1  Attention Deficit Disorder with Hyperactivity/ (34740)
2  ((attention* or behav*) adj3 (defic* or disorder* or dysfunc*)).tw. (77679)
3  adhd.tw. (31201)
4  "ad/hd".tw. (387)
5  addh.tw. (122)
6  adhs.tw. (643)
7  or/1-6 (91163)
8  exp Child/ (2154627)
9  Adolescent/ (2217598)
10  (child* or boy* or girl* or toddler* or infant* or pre?school* or schoolchild* or schoolgirl* or schoolboy* or adolescen* or juvenile* or teen* or young* person* or young* people* or youth* or minor?).tw. (2637404)
11  Minors/ (2824)
12  Pediatrics/ (57913)
13  (pediatric* or paediatric*).tw. (444882)
14  or/8-13 (4595440)
15  (stress* adj2 (event* or generat* or dependent or minor or major)).tw. (27009)
16  oxidative.tw. (394325)
17  15 not 16 (22312)
18  daily hassle?.tw. (660)
19  social structure/ (38)
20  social skills/ (2685)
21  Social Support/ (78822)
22  Social Integration/ (190)
23  exp Social Isolation/ (25590)
24  social capital/ (1645)
25  (social adj (thinning or structure or architecture or group? or experience? or skill? or support or ratings or status or integrat* or relationship? or exclusion or contact? or dysfunction* or depriv* or isolat* or capital)).tw. (109929)
26  SNA.tw. (3639)
27  personal network.tw. (233)
28  (social adj2 process*).tw. (8055)
29  socialising.tw. (336)
30  Interpersonal Relations/ (77080)
31  ((interpersonal or affiliative or supportive) adj1 relationship?).tw. (7170)
32  (shared adj (understanding or norms or values or identit*)).tw. (2101)
33  (community adj (participation or inclusion or involve*)).tw. (5888)
34  "Involvement in a life situation".tw. (14)
35  participation.tw. (183559)
36  ((formal or intimate or close) adj relationships).tw. (5333)
37  Friends/ (6786)
38  friend*.tw. (127175)
39  ((rejected or excluded or disliked) adj children).tw. (341)
40  ((peer? or inter?personal) adj2 (accept* or problem? or conflict or likability or liked or status or interact* or function* or relationship? or interaction? or reject*)).tw. (19755)
41  ((number? or quality or quantity) adj2 relationship?).tw. (7446)
42  Bullying/ (6410)
43  (victim* or bullied or bully*).tw. (69498)
44  marginali#ed.tw. (8978)
45  Loneliness/ (6305)
46  lonel*.tw. (13235)
47  (popular* or unpopular*).tw. (139680)
48  social network analysis/ (265)
49  social network.tw. (12528)
50  exp Social Networking/ (6031)
51  Social Media/ (15905)
52  (48 or 49) not (50 or 51) (10294)
53  or/17-47,52 (793900)
54  7 and 14 and 53 (3962)
55  animal/ (7303730)
56  human/ (21387167)
57  55 not (55 and 56) (5107643)
58  54 not 57 (3880)
59  limit 58 to english language (3592)
60  limit 59 to "remove preprint records" (3589)
61  "Systematic Review"/ or "Review"/ (3293800)
62  exp clinical trial/ (975637)
63  exp qualitative research/ (82620)
64  or/61-63 (4338017)
65  60 not 64 (2840)

**Source: CINAHL**
Interface: EBSCOhost
Database coverage dates: 1981 to present
Search date: 31 July 2023
Retrieved records: 1,510
Search strategy:

S1	(MH "Attention Deficit Hyperactivity Disorder") (18,960)

S2	TI ( ((attention* or behav*) n2 (defic* or disorder* or dysfunc*)) ) OR AB ( ((attention* or behav*) n2 (defic* or disorder* or dysfunc*)) ) (25,787)

S3	TI adhd OR AB adhd (15,315)

S4	TI "ad/hd" OR AB "ad/hd" (109)

S5	TI addh OR AB addh (9)

S6	TI adhs OR AB adhs (549)

S7	S1 OR S2 OR S3 OR S4 OR S5 OR S6 (36,471)

S8	(MH "Child+") (748,784)

S9	(MH "Adolescence") (598,085)

S10	TI ( (child* or boy* or girl* or toddler* or infant* or "pre#school*" or schoolchild* or schoolgirl* or schoolboy* or adolescen* or juvenile* or teen* or "young* person*" or "young* people*" or youth* or minor#) ) OR AB ( (child* or boy* or girl* or toddler* or infant* or "pre#school*" or schoolchild* or schoolgirl* or schoolboy* or adolescen* or juvenile* or teen* or "young* person*" or "young* people*" or youth* or minor#) ) (887,514)

S11	(MH "Minors (Legal)") (807)

S12	(MH "Pediatrics") (21,045)

S13	TI ( (pediatric* or paediatric*) ) OR AB ( (pediatric* or paediatric*) ) (172,808)

S14	S8 OR S9 OR S10 OR S11 OR S12 OR S13 (1,424,266)

S15	TI ( (stress* n1 (event* or generat* or dependent or minor or major)) ) OR AB ( (stress* n1 (event* or generat* or dependent or minor or major)) ) (5,981)

S16	TI oxidative OR AB oxidative (32,689)

S17	S15 NOT S16 (5,684)

S18	TI "daily hassle#" OR AB "daily hassle#" (369)

S19	(MH "Social Structure+") (578)

S20	(MH "Social Skills") (3,798)

S21	(MH "Support, Social") (3,492)

S22	(MH "Social Integration") (284)

S23	(MH "Social Isolation+") (17,390)

S24	(MH "Social Capital") (3,010)

S25	TI ( (social n (thinning or structure or architecture or group# or experience# or skill# or support or ratings or status or integrat* or relationship# or exclusion or contact# or dysfunction* or depriv* or isolat* or capital)) ) OR AB ( (social n (thinning or structure or architecture or group# or experience# or skill# or support or ratings or status or integrat* or relationship# or exclusion or contact# or dysfunction* or depriv* or isolat* or capital)) ) (704)

S26	TI SNA OR AB SNA (677)

S27	TI "personal network" OR AB "personal network" (124)

S28	TI (social n1 process*) OR AB (social n1 process*) (3,720)

S29	TI socialising OR AB socialising (274)

S30	(MH "Interpersonal Relations") (58,166)

S31	TI ( ((interpersonal or affiliative or supportive) n relationship#) ) OR AB ( ((interpersonal or affiliative or supportive) n relationship#) ) (24)

S32	TI ( (shared n (understanding or norms or values or identit*)) ) OR AB ( (shared n (understanding or norms or values or identit*)) ) (7)

S33	TI ( (community n (participation or inclusion or involve*)) ) OR AB ( (community n (participation or inclusion or involve*)) ) (63)

S34	TI "Involvement in a life situation" OR AB "Involvement in a life situation" (9)

S35	TI participation OR AB participation (84,451)

S36	TI ( ((formal or intimate or close) n relationships) ) OR AB ( ((formal or intimate or close) n relationships) ) (29)

S37	(MH "Friendship") (7,624)

S38	TI friend* OR AB friend* (44,279)

S39	TI ( ((rejected or excluded or disliked) n children) ) OR AB ( ((rejected or excluded or disliked) n children) ) (24)

S40	TI ( ((peer# or inter#personal) n1 (accept* or problem# or conflict or likability or liked or status or interact* or function* or relationship# or interaction# or reject*)) ) OR AB ( ((peer# or inter#personal) n1 (accept* or problem# or conflict or likability or liked or status or interact* or function* or relationship# or interaction# or reject*)) ) (11,325)

S41	TI ( ((number# or quality or quantity) n1 relationship#) ) OR AB ( ((number# or quality or quantity) n1 relationship#) ) (4,447)

S42	(MH "Bullying") (9,428)

S43	TI ( (victim* or bullied or bully*) ) OR AB ( (victim* or bullied or bully*) ) (37,319)

S44	TI marginali?ed OR AB marginali?ed (5,763)

S45	TI lonel* OR AB lonel* (8,533)

S46	TI ( (popular* or unpopular*) ) OR AB ( (popular* or unpopular*) ) (35,038)

S47	(MH "Social Networks") (15,229)

S48	(MH "Social Network Analysis") (203)

S49	TI "social network" OR AB "social network" (6,631)

S50	S47 OR S48 OR S49 (19,464)

S51	(MH "Social Media+") (22,158)

S52	(MH "Online Social Networking") (710)

S53	S51 OR S52 (22,511)

S54	S50 NOT S53 (18,378)

S55	S17 OR S18 OR S19 OR S20 OR S21 OR S22 OR S23 OR S24 OR S25 OR S26 OR S27 OR S28 OR S29 OR S30 OR S31 OR S32 OR S33 OR S34 OR S35 OR S36 OR S37 OR S38 OR S39 OR S40 OR S41 OR S42 OR S43 OR S44 OR S45 OR S46 OR S54 (309,288)

S56	S7 AND S14 AND S55 (1,777)

S57	(MH "Animals") (97,152)

S58	(MH "Human") (2,680,083)

S59	S57 NOT (S57 AND S58) (88,785)

S60	S56 NOT S59 (1,775)

S61	S56 NOT S59 Limiters - English Language (1,742)

S62	(MH "Literature Review+") (138,352)

S63	(MH "Clinical Trials+") (348,942)

S64	(MH "Qualitative Studies+") (182,499)

S65	S62 OR S63 OR S64 (651,237)

S66	S61 NOT S65 (1,510)

##### **Source: APA PsycINFO**

Interface: EBSCOhost

Database coverage dates: 1597 to present

Search date: 31 July 2023

Retrieved records: 4,217

Search strategy:

S1	DE "Attention Deficit Disorder with Hyperactivity" (34,540)

S2	TI ( ((attention* or behav*) n2 (defic* or disorder* or dysfunc*)) ) OR AB ( ((attention* or behav*) n2 (defic* or disorder* or dysfunc*)) ) (70,817)

S3	TI adhd OR AB adhd (31,987)

S4	TI "ad/hd" OR AB "ad/hd" (580)

S5	TI addh OR AB addh (130)

S6	TI adhs OR AB adhs (681)

S7	S1 OR S2 OR S3 OR S4 OR S5 OR S6 (81,846)

S8	AG Childhood (604,843)

S9	AG Adolescence (499,656)

S10	TI ( (child* or boy* or girl* or toddler* or infant* or pre#school* or schoolchild* or schoolgirl* or schoolboy* or adolescen* or juvenile* or teen* or "young* person*" or "young* people*" or youth* or minor#) ) OR AB ( (child* or boy* or girl* or toddler* or infant* or pre#school* or schoolchild* or schoolgirl* or schoolboy* or adolescen* or juvenile* or teen* or "young* person*" or "young* people*" or youth* or minor#) ) (1,122,403)

S11	DE "Pediatrics" (32,896)

S12	TI ( (pediatric* or paediatric*) ) OR AB ( (pediatric* or paediatric*) ) (43,279)

S13	S8 OR S9 OR S10 OR S11 OR S12 (1,354,967)

S14	TI ( (stress* n1 (event* or generat* or dependent or minor or major)) ) OR AB ( (stress* n1 (event* or generat* or dependent or minor or major)) ) (15,143)

S15	TI oxidative OR AB oxidative (12,591)

S16	S14 NOT S15 (14,950)

S17	TI daily hassle# OR AB daily hassle# (1,247)

S18	DE "Social Structure" (6,494)

S19	DE "Social Skills" (16,487)

S20	DE "Social Support" (66,474)

S21	DE "Social Exclusion" (2,263)

S22	DE "Social Deprivation" (1,016)

S23	DE "Social Isolation" (13,596)

S24	DE "Social Capital" (7,641)

S25	TI ( (social n (thinning or structure or architecture or group# or experience# or skill# or support or ratings or status or integrat* or relationship# or exclusion or contact# or dysfunction* or depriv* or isolat* or capital)) ) OR AB ( (social n (thinning or structure or architecture or group# or experience# or skill# or support or ratings or status or integrat* or relationship# or exclusion or contact# or dysfunction* or depriv* or isolat* or capital)) ) (1,337)

S26	TI SNA OR AB SNA (627)

S27	TI "personal network" OR AB "personal network" (330)

S28	TI (social n1 process*) OR AB (social n1 process*) (17,352)

S29	TI socialising OR AB socialising (294)

S30	DE "Interpersonal Relationships" (23,831)

S31	TI ( ((interpersonal or affiliative or supportive) n relationship#) ) OR AB ( ((interpersonal or affiliative or supportive) n relationship#) ) (46)

S32	TI ( (shared n (understanding or norms or values or identit*)) ) OR AB ( (shared n (understanding or norms or values or identit*)) ) (8)

S33	DE "Community Involvement" (6,618)

S34	TI ( (community n (participation or inclusion or involve*)) ) OR AB ( (community n (participation or inclusion or involve*)) ) (77)

S35	TI "Involvement in a life situation" OR AB "Involvement in a life situation" (13)

S36	TI participation OR AB participation (111,017)

S37	DE "Close Relationships" (509)

S38	TI ( ((formal or intimate or close) n relationships) ) OR AB ( ((formal or intimate or close) n relationships) ) (77)

S39	DE "Friendship" (10,988)

S40	TI friend* OR AB friend* (77,798)

S41	TI ( ((rejected or excluded or disliked) n children) ) OR AB ( ((rejected or excluded or disliked) n children) ) (16)

S42	DE "Peer Relations" (18,437)

S43	DE "Interpersonal Interaction" (33,565)

S44	TI ( ((peer# or inter#personal) n1 (accept* or problem# or conflict or likability or liked or status or interact* or function* or relationship# or interaction# or reject*)) ) OR AB ( ((peer# or inter#personal) n1 (accept* or problem# or conflict or likability or liked or status or interact* or function* or relationship# or interaction# or reject*)) ) (42,252)

S45	DE "Relationship Quality" (7,402)

S46	TI ( ((number# or quality or quantity) n1 relationship#) ) OR AB ( ((number# or quality or quantity) n1 relationship#) ) (10,995)

S47	DE "Bullying" (11,518)

S48	DE "Victimization" (25,710)

S49	TI ( (victim* or bullied or bully*) ) OR AB ( (victim* or bullied or bully*) ) (77,724)

S50	TI marginali?ed OR AB marginali?ed (12,487)

S51	DE "Loneliness" (7,620)

S52	TI lonel* OR AB lonel* (14,683)

S53	DE "Popularity" (1,970)

S54	TI ( (popular* or unpopular*) ) OR AB ( (popular* or unpopular*) ) (60,316)

S55	DE "Social Networks" (14,523)

S56	DE "Social Network Analysis" (1,761)

S57	TI "social network" OR AB "social network" (14,613)

S58	S55 OR S56 OR S57 (23,453)

S59	DE "Social Media" (16,623)

S60	DE "Online Social Networks" (9,849)

S61	S59 OR S60 (23,383)

S62	S58 NOT S61 (20,841)

S63	S16 OR S17 OR S18 OR S19 OR S20 OR S21 OR S22 OR S23 OR S24 OR S25 OR S26 OR S27 OR S28 OR S29 OR S30 OR S31 OR S32 OR S33 OR S34 OR S35 OR S36 OR S37 OR S38 OR S39 OR S40 OR S41 OR S42 OR S43 OR S44 OR S45 OR S46 OR S47 OR S48 OR S49 OR S50 OR S51 OR S52 OR S53 OR S54 OR S62 (559,487)

S64	S7 AND S13 AND S63 (4,566)

S65	DE "Animals" (267,530)

S66	S64 NOT S65 (4,529)

S67	DE "Literature Review" OR DE "Systematic Review" (23,809)

S68	DE "Clinical Trials" OR DE "Randomized Controlled Trials" OR DE "Randomized Clinical Trials" (13,656)

S69	DE "Qualitative Methods" OR DE "Focus Group" OR DE "Grounded Theory" OR DE "Interpretative Phenomenological Analysis" OR DE "Narrative Analysis" OR DE "Semi-Structured Interview" OR DE "Thematic Analysis" OR DE "Focus Group Interview" (21,526)

S70	S67 OR S68 OR S69 (58,752)

S71	S66 NOT S70 (4,489)

S72	S66 NOT S70 Limiters – English (4,217)

First update, 30 January

**Source: Embase 1947-Present, updated daily**
Interface: OvidSP

Database coverage dates: 1947 to present

Search date: 30 January 2024

Retrieved records: 261

Search strategy:

1 attention deficit hyperactivity disorder/ (12394)

2 ((attention* or behav*) adj3 (defic* or disorder* or dysfunc*)).tw. (109915)

3 adhd.tw. (46992)

4 "ad/hd".tw. (507)

5 addh.tw. (150)

6 adhs.tw. (727)

7 or/1-6 (127960)

8 exp child/ (3573305)

9 adolescent/ (1957188)

10 (child* or boy* or girl* or toddler* or infant* or pre?school* or schoolchild* or schoolgirl* or schoolboy* or adolescen* or juvenile* or teen* or young* person* or young* people* or youth* or minor?).tw. (3667845)

11 "minor (person)"/ (976)

12 exp pediatrics/ (142300)

13 (pediatric* or paediatric*).tw. (740280)

14 or/8-13 (5800391)

15 (stress* adj2 (event* or generat* or dependent or minor or major)).tw. (34628)

16 oxidative.tw. (513016)

17 15 not 16 (28534)

18 daily hassle?.tw. (792)

19 social structure/ (6405)

20 social competence/ (8195)

21 social support/ (118174)

22 integration/ (7546)

23 exp social isolation/ (35549)

24 social capital/ (4131)

25 (social adj (thinning or structure or architecture or group? or experience? or skill? or support or ratings or status or integrat* or relationship? or exclusion or contact? or dysfunction* or depriv* or isolat* or capital)).tw. (141419)

26 SNA.tw. (5057)

27 personal network.tw. (264)

28 (social adj2 process*).tw. (9458)

29 socialising.tw. (513)

30 ((interpersonal or affiliative or supportive) adj1 relationship?).tw. (10011)

31 (shared adj (understanding or norms or values or identit*)).tw. (2508)

32 community participation/ (4533)

33 (community adj (participation or inclusion or involve*)).tw. (7052)

34 "Involvement in a life situation".tw. (20)

35 participation.tw. (253732)

36 ((formal or intimate or close) adj relationships).tw. (6707)

37 friendship/ (4486)

38 friend*.tw. (162890)

39 ((rejected or excluded or disliked) adj children).tw. (716)

40 ((peer? or inter?personal) adj2 (accept* or problem? or conflict or likability or liked or status or interact* or function* or relationship? or interaction? or reject*)).tw. (25880)

41 ((number? or quality or quantity) adj2 relationship?).tw. (9529)

42 bullying/ (10842)

43 (victim* or bullied or bully*).tw. (89734)

44 marginali#ed.tw. (11025)

45 loneliness/ (15509)

46 lonel*.tw. (17073)

47 (popular* or unpopular*).tw. (179791)

48 social network/ or social network analysis/ (25678)

49 social network.tw. (14778)

50 online social network/ or social media/ (52956)

51 (48 or 49) not 50 (28055)

52 or/17-47,51 (998123)

53 7 and 14 and 52 (5548)

54 animal/ (2149774)

55 human/ (27321255)

56 54 not (54 and 55) (1629936)

57 53 not 56 (5540)

58 limit 57 to english language (5175)

59 limit 58 to "remove preprint records" (5162)

60 exp "review"/ (3272014)

61 exp clinical trial/ (1899211)

62 exp qualitative research/ (123943)

63 or/60-62 (5093510)

64 59 not 63 (4160)

65 limit 64 to dc=20230731-20240130 (261)

**Source: Ovid MEDLINE(R) ALL**
Interface OvidSP

Database coverage dates: 1946 to 29 January 2024

Search date: 30 January 2024

Retrieved records: 113

Search strategy:

1 Attention Deficit Disorder with Hyperactivity/ (35465)

2 ((attention* or behav*) adj3 (defic* or disorder* or dysfunc*)).tw. (80149)

3 adhd.tw. (32367)

4 "ad/hd".tw. (395)

5 addh.tw. (123)

6 adhs.tw. (662)

7 or/1-6 (93948)

8 exp Child/ (2185624)

9 Adolescent/ (2233882)

10 (child* or boy* or girl* or toddler* or infant* or pre?school* or schoolchild* or schoolgirl* or schoolboy* or adolescen* or juvenile* or teen* or young* person* or young* people* or youth* or minor?).tw. (2700147)

11 Minors/ (2840)

12 Pediatrics/ (58001)

13 (pediatric* or paediatric*).tw. (461150)

14 or/8-13 (4668858)

15 (stress* adj2 (event* or generat* or dependent or minor or major)).tw. (27834)

16 oxidative.tw. (409946)

17 15 not 16 (22993)

18 daily hassle?.tw. (668)

19 social structure/ (47)

20 social skills/ (2754)

21 Social Support/ (79517)

22 Social Integration/ (203)

23 exp Social Isolation/ (26162)

24 social capital/ (1706)

25 (social adj (thinning or structure or architecture or group? or experience? or skill? or support or ratings or status or integrat* or relationship? or exclusion or contact? or dysfunction* or depriv* or isolat* or capital)).tw. (114414)

26 SNA.tw. (3753)

27 personal network.tw. (239)

28 (social adj2 process*).tw. (8330)

29 socialising.tw. (357)

30 Interpersonal Relations/ (77515)

31 ((interpersonal or affiliative or supportive) adj1 relationship?).tw. (7508)

32 (shared adj (understanding or norms or values or identit*)).tw. (2232)

33 (community adj (participation or inclusion or involve*)).tw. (6085)

34 "Involvement in a life situation".tw. (14)

35 participation.tw. (189792)

36 ((formal or intimate or close) adj relationships).tw. (5525)

37 Friends/ (6986)

38 friend*.tw. (134954)

39 ((rejected or excluded or disliked) adj children).tw. (349)

40 ((peer? or inter?personal) adj2 (accept* or problem? or conflict or likability or liked or status or interact* or function* or relationship? or interaction? or reject*)).tw. (20543)

41 ((number? or quality or quantity) adj2 relationship?).tw. (7713)

42 Bullying/ (6643)

43 (victim* or bullied or bully*).tw. (71403)

44 marginali#ed.tw. (9799)

45 Loneliness/ (6682)

46 lonel*.tw. (14254)

47 (popular* or unpopular*).tw. (146246)

48 social network analysis/ (290)

49 social network.tw. (13047)

50 exp Social Networking/ (6187)

51 Social Media/ (16793)

52 (48 or 49) not (50 or 51) (10719)

53 or/17-47,52 (823650)

54 7 and 14 and 53 (4073)

55 animal/ (7382349)

56 human/ (21747540)

57 55 not (55 and 56) (5157999)

58 54 not 57 (3989)

59 limit 58 to english language (3700)

60 limit 59 to "remove preprint records" (3696)

61 "Systematic Review"/ or "Review"/ (3388883)

62 exp clinical trial/ (988137)

63 exp qualitative research/ (85643)

64 or/61-63 (4448296)

65 60 not 64 (2929)

66 ("202308*" or "202309*" or "20231*" or 2024*).dt,ez,ed. (915936)

67 65 and 66 (113)

**Source: CINAHL**
Interface: EBSCOhost
Database coverage dates: 1981 to present
Search date: 30 January 2024
Retrieved records: 48
Search strategy:

S1	(MH "Attention Deficit Hyperactivity Disorder")	19,326

S2	TI ( ((attention* or behav*) n2 (defic* or disorder* or dysfunc*)) ) OR AB ( ((attention* or behav*) n2 (defic* or disorder* or dysfunc*)) )	26,556

S3	TI adhd OR AB adhd	15,862

S4	TI "ad/hd" OR AB "ad/hd"	110

S5	TI addh OR AB addh	9

S6	TI adhs OR AB adhs	694

S7	S1 OR S2 OR S3 OR S4 OR S5 OR S6	37,722

S8	(MH "Child+")	760,008

S9	(MH "Adolescence")	609,602

S10	TI ( (child* or boy* or girl* or toddler* or infant* or "pre#school*" or schoolchild* or schoolgirl* or schoolboy* or adolescen* or juvenile* or teen* or "young* person*" or "young* people*" or youth* or minor#) ) OR AB ( (child* or boy* or girl* or toddler* or infant* or "pre#school*" or schoolchild* or schoolgirl* or schoolboy* or adolescen* or juvenile* or teen* or "young* person*" or "young* people*" or youth* or minor#) )	910,767

S11	(MH "Minors (Legal)")	814

S12	(MH "Pediatrics")	21,286

S13	TI ( (pediatric* or paediatric*) ) OR AB ( (pediatric* or paediatric*) )	178,058

S14	S8 OR S9 OR S10 OR S11 OR S12 OR S13	1,455,711

S15	TI ( (stress* n1 (event* or generat* or dependent or minor or major)) ) OR AB ( (stress* n1 (event* or generat* or dependent or minor or major)) )	6,113

S16	TI oxidative OR AB oxidative	33,687

S17	S15 NOT S16	5,811

S18	TI "daily hassle#" OR AB "daily hassle#"	372

S19	(MH "Social Structure+")	746

S20	(MH "Social Skills")	3,975

S21	(MH "Support, Social")	5,001

S22	(MH "Social Integration")	341

S23	(MH "Social Isolation+")	18,451

S24	(MH "Social Capital")	3,109

S25	TI ( (social n (thinning or structure or architecture or group# or experience# or skill# or support or ratings or status or integrat* or relationship# or exclusion or contact# or dysfunction* or depriv* or isolat* or capital)) ) OR AB ( (social n (thinning or structure or architecture or group# or experience# or skill# or support or ratings or status or integrat* or relationship# or exclusion or contact# or dysfunction* or depriv* or isolat* or capital)) )	736

S26	TI SNA OR AB SNA	694

S27	TI "personal network" OR AB "personal network"	126

S28	TI (social n1 process*) OR AB (social n1 process*)	3,788

S29	TI socialising OR AB socialising	279

S30	(MH "Interpersonal Relations")	60,254

S31	TI ( ((interpersonal or affiliative or supportive) n relationship#) ) OR AB ( ((interpersonal or affiliative or supportive) n relationship#) )	24

S32	TI ( (shared n (understanding or norms or values or identit*)) ) OR AB ( (shared n (understanding or norms or values or identit*)) )	8

S33	TI ( (community n (participation or inclusion or involve*)) ) OR AB ( (community n (participation or inclusion or involve*)) )	64

S34	TI "Involvement in a life situation" OR AB "Involvement in a life situation"	9

S35	TI participation OR AB participation	86,621

S36	TI ( ((formal or intimate or close) n relationships) ) OR AB ( ((formal or intimate or close) n relationships) )	33

S37	(MH "Friendship")	7,882

S38	TI friend* OR AB friend*	45,358

S39	TI ( ((rejected or excluded or disliked) n children) ) OR AB ( ((rejected or excluded or disliked) n children) )	25

S40	TI ( ((peer# or inter#personal) n1 (accept* or problem# or conflict or likability or liked or status or interact* or function* or relationship# or interaction# or reject*)) ) OR AB ( ((peer# or inter#personal) n1 (accept* or problem# or conflict or likability or liked or status or interact* or function* or relationship# or interaction# or reject*)) )	11,670

S41	TI ( ((number# or quality or quantity) n1 relationship#) ) OR AB ( ((number# or quality or quantity) n1 relationship#) )	4,552

S42	(MH "Bullying")	9,710

S43	TI ( (victim* or bullied or bully*) ) OR AB ( (victim* or bullied or bully*) )	38,356

S44	TI marginali?ed OR AB marginali?ed	6,130

S45	TI lonel* OR AB lonel*	9,005

S46	TI ( (popular* or unpopular*) ) OR AB ( (popular* or unpopular*) )	35,646

S47	(MH "Social Networks")	15,461

S48	(MH "Social Network Analysis")	280

S49	TI "social network" OR AB "social network"	6,761

S50	S47 OR S48 OR S49	19,793

S51	(MH "Social Media+")	23,492

S52	(MH "Online Social Networking")	790

S53	S51 OR S52	23,889

S54	S50 NOT S53	18,672

S55	S17 OR S18 OR S19 OR S20 OR S21 OR S22 OR S23 OR S24 OR S25 OR S26 OR S27 OR S28 OR S29 OR S30 OR S31 OR S32 OR S33 OR S34 OR S35 OR S36 OR S37 OR S38 OR S39 OR S40 OR S41 OR S42 OR S43 OR S44 OR S45 OR S46 OR S54	318,974

S56	S7 AND S14 AND S55	1,845

S57	(MH "Animals")	96,578

S58	(MH "Human")	2,759,496

S59	S57 NOT (S57 AND S58)	87,990

S60	S56 NOT S59	1,843

S61	S56 NOT S59 - Limiters - English Language	1,809

S62	(MH "Literature Review+")	148,484

S63	(MH "Clinical Trials+")	354,159

S64	(MH "Qualitative Studies+")	189,949

S65	S62 OR S63 OR S64	673,495

S66	S61 NOT S65	1,567

S67	EM 20230801-20240130	127,144

S68	S66 AND S67	48

**Source: APA PsycINFO**

Interface: EBSCOhost

Database coverage dates: 1597 to present

Search date: 30 January 2024

Retrieved records: 111

Search strategy:

S1	DE "Attention Deficit Disorder with Hyperactivity"	35,376

S2	TI ( ((attention* or behav*) n2 (defic* or disorder* or dysfunc*)) ) OR AB ( ((attention* or behav*) n2 (defic* or disorder* or dysfunc*)) )	72,338

S3	TI adhd OR AB adhd	32,818

S4	TI "ad/hd" OR AB "ad/hd"	585

S5	TI addh OR AB addh	130

S6	TI adhs OR AB adhs	692

S7	S1 OR S2 OR S3 OR S4 OR S5 OR S6	83,613

S8	AG Childhood	615,978

S9	AG Adolescence	511,049

S10	TI ( (child* or boy* or girl* or toddler* or infant* or pre#school* or schoolchild* or schoolgirl* or schoolboy* or adolescen* or juvenile* or teen* or "young* person*" or "young* people*" or youth* or minor#) ) OR AB ( (child* or boy* or girl* or toddler* or infant* or pre#school* or schoolchild* or schoolgirl* or schoolboy* or adolescen* or juvenile* or teen* or "young* person*" or "young* people*" or youth* or minor#) )	1,143,736

S11	DE "Pediatrics"	33,962

S12	TI ( (pediatric* or paediatric*) ) OR AB ( (pediatric* or paediatric*) )	44,524

S13	S8 OR S9 OR S10 OR S11 OR S12	1,380,813

S14	TI ( (stress* n1 (event* or generat* or dependent or minor or major)) ) OR AB ( (stress* n1 (event* or generat* or dependent or minor or major)) )	15,480

S15	TI oxidative OR AB oxidative	12,925

S16	S14 NOT S15	15,284

S17	TI daily hassle# OR AB daily hassle#	1,253

S18	DE "Social Structure"	6,541

S19	DE "Social Skills"	16,722

S20	DE "Social Support"	67,796

S21	DE "Social Exclusion"	2,341

S22	DE "Social Deprivation"	1,031

S23	DE "Social Isolation"	13,915

S24	DE "Social Capital"	7,868

S25	TI ( (social n (thinning or structure or architecture or group# or experience# or skill# or support or ratings or status or integrat* or relationship# or exclusion or contact# or dysfunction* or depriv* or isolat* or capital)) ) OR AB ( (social n (thinning or structure or architecture or group# or experience# or skill# or support or ratings or status or integrat* or relationship# or exclusion or contact# or dysfunction* or depriv* or isolat* or capital)) )	1,381

S26	TI SNA OR AB SNA	651

S27	TI "personal network" OR AB "personal network"	335

S28	TI (social n1 process*) OR AB (social n1 process*)	17,708

S29	TI socialising OR AB socialising	321

S30	DE "Interpersonal Relationships"	25,666

S31	TI ( ((interpersonal or affiliative or supportive) n relationship#) ) OR AB ( ((interpersonal or affiliative or supportive) n relationship#) )	48

S32	TI ( (shared n (understanding or norms or values or identit*)) ) OR AB ( (shared n (understanding or norms or values or identit*)) )	9

S33	DE "Community Involvement"	6,911

S34	TI ( (community n (participation or inclusion or involve*)) ) OR AB ( (community n (participation or inclusion or involve*)) )	82

S35	TI "Involvement in a life situation" OR AB "Involvement in a life situation"	13

S36	TI participation OR AB participation	113,983

S37	DE "Close Relationships"	720

S38	TI ( ((formal or intimate or close) n relationships) ) OR AB ( ((formal or intimate or close) n relationships) )	85

S39	DE "Friendship"	11,202

S40	TI friend* OR AB friend*	79,602

S41	TI ( ((rejected or excluded or disliked) n children) ) OR AB ( ((rejected or excluded or disliked) n children) )	16

S42	DE "Peer Relations"	18,754

S43	DE "Interpersonal Interaction"	33,791

S44	TI ( ((peer# or inter#personal) n1 (accept* or problem# or conflict or likability or liked or status or interact* or function* or relationship# or interaction# or reject*)) ) OR AB ( ((peer# or inter#personal) n1 (accept* or problem# or conflict or likability or liked or status or interact* or function* or relationship# or interaction# or reject*)) )	43,201

S45	DE "Relationship Quality"	7,685

S46	TI ( ((number# or quality or quantity) n1 relationship#) ) OR AB ( ((number# or quality or quantity) n1 relationship#) )	11,329

S47	DE "Bullying"	11,908

S48	DE "Victimization"	26,408

S49	TI ( (victim* or bullied or bully*) ) OR AB ( (victim* or bullied or bully*) )	79,694

S50	TI marginali?ed OR AB marginali?ed	13,315

S51	DE "Loneliness"	8,131

S52	TI lonel* OR AB lonel*	15,470

S53	DE "Popularity"	2,033

S54	TI ( (popular* or unpopular*) ) OR AB ( (popular* or unpopular*) )	61,858

S55	DE "Social Networks"	14,928

S56	DE "Social Network Analysis"	1,858

S57	TI "social network" OR AB "social network"	15,045

S58	S55 OR S56 OR S57	24,106

S59	DE "Social Media"	18,260

S60	DE "Online Social Networks"	10,141

S61	S59 OR S60	25,141

S62	S58 NOT S61	21,389

S63	S16 OR S17 OR S18 OR S19 OR S20 OR S21 OR S22 OR S23 OR S24 OR S25 OR S26 OR S27 OR S28 OR S29 OR S30 OR S31 OR S32 OR S33 OR S34 OR S35 OR S36 OR S37 OR S38 OR S39 OR S40 OR S41 OR S42 OR S43 OR S44 OR S45 OR S46 OR S47 OR S48 OR S49 OR S50 OR S51 OR S52 OR S53 OR S54 OR S62	574,261

S64	S7 AND S13 AND S63	4,689

S65	DE "Animals"	268,344

S66	S64 NOT S65	4,651

S67	DE "Literature Review" OR DE "Systematic Review"	23,869

S68	DE "Clinical Trials" OR DE "Randomized Controlled Trials" OR DE "Randomized Clinical Trials"	13,812

S69	DE "Qualitative Methods" OR DE "Focus Group" OR DE "Grounded Theory" OR DE "Interpretative Phenomenological Analysis" OR DE "Narrative Analysis" OR DE "Semi-Structured Interview" OR DE "Thematic Analysis" OR DE "Focus Group Interview"	22,212

S70	S67 OR S68 OR S69	59,652

S71	S66 NOT S70	4,611

S72	S66 NOT S70 - Limiters - English language	4,334

S73	RD 20230801-20240130	97,365

S74	S72 AND S73	111

Second update, 23 April 2025

**Source: Embase 1947-Present, updated daily**
Interface: OvidSP

Database coverage dates: 1947 to present

Search date: 23 April 2025

Retrieved records: 579

Search strategy:

1	attention deficit hyperactivity disorder/	88277

2	((attention* or behav*) adj3 (defic* or disorder* or dysfunc*)).tw.	118189

3	adhd.tw.	51041

4	"ad/hd".tw.	514

5	addh.tw.	156

6	adhs.tw.	747

7	or/1-6	159354

8	exp child/	3806216

9	adolescent/	2083416

10	(child* or boy* or girl* or toddler* or infant* or pre?school* or schoolchild* or schoolgirl* or schoolboy* or adolescen* or juvenile* or teen* or young* person* or young* people* or youth* or minor?).tw.	3924149

11	"minor (person)"/	1082

12	exp pediatrics/	151208

13	(pediatric* or paediatric*).tw.	805924

14	or/8-13	6189764

15	(stress* adj2 (event* or generat* or dependent or minor or major)).tw.	36933

16	oxidative.tw.	559089

17	15 not 16	30451

18	daily hassle?.tw.	815

19	social structure/	6714

20	social competence/	9583

21	social support/	129802

22	integration/	8477

23	exp social isolation/	38880

24	social capital/	4557

25	(social adj (thinning or structure or architecture or group? or experience? or skill? or support or ratings or status or integrat* or relationship? or exclusion or contact? or dysfunction* or depriv* or isolat* or capital)).tw.	154900

26	SNA.tw.	5447

27	personal network.tw.	283

28	(social adj2 process*).tw.	10248

29	socialising.tw.	586

30	((interpersonal or affiliative or supportive) adj1 relationship?).tw.	11224

31	(shared adj (understanding or norms or values or identit*)).tw.	2864

32	community participation/	5222

33	(community adj (participation or inclusion or involve*)).tw.	7725

34	"Involvement in a life situation".tw.	20

35	participation.tw.	273732

36	((formal or intimate or close) adj relationships).tw.	7222

37	friendship/	4866

38	friend*.tw.	185361

39	((rejected or excluded or disliked) adj children).tw.	768

40	((peer? or inter?personal) adj2 (accept* or problem? or conflict or likability or liked or status or interact* or function* or relationship? or interaction? or reject*)).tw.	28424

41	((number? or quality or quantity) adj2 relationship?).tw.	10263

42	bullying/	12064

43	(victim* or bullied or bully*).tw.	95201

44	marginali#ed.tw.	13457

45	loneliness/	18267

46	lonel*.tw.	20004

47	(popular* or unpopular*).tw.	196638

48	social network/ or social network analysis/	29070

49	social network.tw.	16057

50	online social network/ or social media/	64595

51	(48 or 49) not 50	31103

52	or/17-47,51	1092414

53	7 and 14 and 52	7203

54	animal/	2187096

55	human/	28990790

56	54 not (54 and 55)	1655155

57	53 not 56	7193

58	limit 57 to english language	6783

59	limit 58 to "remove preprint records"	6758

60	exp "review"/	3480499

61	exp clinical trial/	2022626

62	exp qualitative research/	145949

63	or/60-62	5435799

64	59 not 63	5400

65	limit 64 to dc=20240130-20250423	579

**Source: Ovid MEDLINE(R) ALL**
Interface OvidSP

Database coverage dates: 1946 to 21 April 2025

Search date: 23 April 2025

Retrieved records: 299

Search strategy:

1	Attention Deficit Disorder with Hyperactivity/	37410

2	((attention* or behav*) adj3 (defic* or disorder* or dysfunc*)).tw.	86620

3	adhd.tw.	35379

4	"ad/hd".tw.	399

5	addh.tw.	127

6	adhs.tw.	695

7	or/1-6	101200

8	exp Child/	2262819

9	Adolescent/	2322038

10	(child* or boy* or girl* or toddler* or infant* or pre?school* or schoolchild* or schoolgirl* or schoolboy* or adolescen* or juvenile* or teen* or young* person* or young* people* or youth* or minor?).tw.	2856537

11	Minors/	2906

12	Pediatrics/	59697

13	(pediatric* or paediatric*).tw.	503149

14	or/8-13	4885155

15	(stress* adj2 (event* or generat* or dependent or minor or major)).tw.	29909

16	oxidative.tw.	451643

17	15 not 16	24707

18	daily hassle?.tw.	686

19	social structure/	50

20	social skills/	3035

21	Social Support/	83975

22	Social Integration/	240

23	exp Social Isolation/	28138

24	social capital/	1887

25	(social adj (thinning or structure or architecture or group? or experience? or skill? or support or ratings or status or integrat* or relationship? or exclusion or contact? or dysfunction* or depriv* or isolat* or capital)).tw.	126501

26	SNA.tw.	4065

27	personal network.tw.	261

28	(social adj2 process*).tw.	9052

29	socialising.tw.	414

30	Interpersonal Relations/	79377

31	((interpersonal or affiliative or supportive) adj1 relationship?).tw.	8406

32	(shared adj (understanding or norms or values or identit*)).tw.	2578

33	(community adj (participation or inclusion or involve*)).tw.	6749

34	"Involvement in a life situation".tw.	14

35	participation.tw.	205799

36	((formal or intimate or close) adj relationships).tw.	6004

37	Friends/	7311

38	friend*.tw.	156877

39	((rejected or excluded or disliked) adj children).tw.	376

40	((peer? or inter?personal) adj2 (accept* or problem? or conflict or likability or liked or status or interact* or function* or relationship? or interaction? or reject*)).tw.	22683

41	((number? or quality or quantity) adj2 relationship?).tw.	8357

42	Bullying/	7289

43	(victim* or bullied or bully*).tw.	76230

44	marginali#ed.tw.	12010

45	Loneliness/	8001

46	lonel*.tw.	17006

47	(popular* or unpopular*).tw.	160977

48	social network analysis/	395

49	social network.tw.	14269

50	exp Social Networking/	6649

51	Social Media/	19537

52	(48 or 49) not (50 or 51)	11691

53	or/17-47,52	901987

54	7 and 14 and 53	4414

55	animal/	7645954

56	human/	22663856

57	55 not (55 and 56)	5295023

58	54 not 57	4324

59	limit 58 to english language	4032

60	limit 59 to "remove preprint records"	4024

61	"Systematic Review"/ or "Review"/	3604003

62	exp clinical trial/	1021121

63	exp qualitative research/	102721

64	or/61-63	4712957

65	60 not 64	3201

66	("2024013*" or "202402*" or "202403*" or "202404*" or "202405*" or "202406*" or "202407*" or "202408*" or "202409*" or "20241*" or 2025*).dt,ez,ed.	2129570

67	65 and 66	299

**Source: CINAHL**
Interface: EBSCOhost
Database coverage dates: 1981 to present
Search date: 23 April 2025
Retrieved records: 114
Search strategy:

S1	(MH "Attention Deficit Hyperactivity Disorder")	20,184

S2	TI ( ((attention* or behav*) n2 (defic* or disorder* or dysfunc*)) ) OR AB ( ((attention* or behav*) n2 (defic* or disorder* or dysfunc*)) )	26,231

S3	TI adhd OR AB adhd	16,168

S4	TI "ad/hd" OR AB "ad/hd"	109

S5	TI addh OR AB addh	9

S6	TI adhs OR AB adhs	670

S7	S1 OR S2 OR S3 OR S4 OR S5 OR S6	39,054

S8	(MH "Child+")	779,621

S9	(MH "Adolescence")	632,140

S10	TI ( (child* or boy* or girl* or toddler* or infant* or "pre#school*" or schoolchild* or schoolgirl* or schoolboy* or adolescen* or juvenile* or teen* or "young* person*" or "young* people*" or youth* or minor#) ) OR AB ( (child* or boy* or girl* or toddler* or infant* or "pre#school*" or schoolchild* or schoolgirl* or schoolboy* or adolescen* or juvenile* or teen* or "young* person*" or "young* people*" or youth* or minor#) )	932,584

S11	(MH "Minors (Legal)")	858

S12	(MH "Pediatrics")	21,352

S13	TI ( (pediatric* or paediatric*) ) OR AB ( (pediatric* or paediatric*) )	184,581

S14	S8 OR S9 OR S10 OR S11 OR S12 OR S13	1,506,407

S15	TI ( (stress* n1 (event* or generat* or dependent or minor or major)) ) OR AB ( (stress* n1 (event* or generat* or dependent or minor or major)) )	6,347

S16	TI oxidative OR AB oxidative	34,252

S17	S15 NOT S16	6,040

S18	TI "daily hassle#" OR AB "daily hassle#"	382

S19	(MH "Social Structure+")	1,178

S20	(MH "Social Skills")	4,397

S21	(MH "Support, Social")	8,950

S22	(MH "Social Integration")	503

S23	(MH "Social Isolation+")	21,114

S24	(MH "Social Capital")	3,395

S25	TI ( (social n (thinning or structure or architecture or group# or experience# or skill# or support or ratings or status or integrat* or relationship# or exclusion or contact# or dysfunction* or depriv* or isolat* or capital)) ) OR AB ( (social n (thinning or structure or architecture or group# or experience# or skill# or support or ratings or status or integrat* or relationship# or exclusion or contact# or dysfunction* or depriv* or isolat* or capital)) )	798

S26	TI SNA OR AB SNA	711

S27	TI "personal network" OR AB "personal network"	131

S28	TI (social n1 process*) OR AB (social n1 process*)	3,997

S29	TI socialising OR AB socialising	298

S30	(MH "Interpersonal Relations")	66,507

S31	TI ( ((interpersonal or affiliative or supportive) n relationship#) ) OR AB ( ((interpersonal or affiliative or supportive) n relationship#) )	26

S32	TI ( (shared n (understanding or norms or values or identit*)) ) OR AB ( (shared n (understanding or norms or values or identit*)) )	9

S33	TI ( (community n (participation or inclusion or involve*)) ) OR AB ( (community n (participation or inclusion or involve*)) )	69

S34	TI "Involvement in a life situation" OR AB "Involvement in a life situation"	9

S35	TI participation OR AB participation	90,065

S36	TI ( ((formal or intimate or close) n relationships) ) OR AB ( ((formal or intimate or close) n relationships) )	37

S37	(MH "Friendship")	8,554

S38	TI friend* OR AB friend*	45,542

S39	TI ( ((rejected or excluded or disliked) n children) ) OR AB ( ((rejected or excluded or disliked) n children) )	26

S40	TI ( ((peer# or inter#personal) n1 (accept* or problem# or conflict or likability or liked or status or interact* or function* or relationship# or interaction# or reject*)) ) OR AB ( ((peer# or inter#personal) n1 (accept* or problem# or conflict or likability or liked or status or interact* or function* or relationship# or interaction# or reject*)) )	12,394

S41	TI ( ((number# or quality or quantity) n1 relationship#) ) OR AB ( ((number# or quality or quantity) n1 relationship#) )	4,862

S42	(MH "Bullying")	10,387

S43	TI ( (victim* or bullied or bully*) ) OR AB ( (victim* or bullied or bully*) )	39,428

S44	TI marginali?ed OR AB marginali?ed	6,947

S45	TI lonel* OR AB lonel*	10,182

S46	TI ( (popular* or unpopular*) ) OR AB ( (popular* or unpopular*) )	35,522

S47	(MH "Social Networks")	16,225

S48	(MH "Social Network Analysis")	460

S49	TI "social network" OR AB "social network"	7,078

S50	S47 OR S48 OR S49	20,825

S51	(MH "Social Media+")	26,535

S52	(MH "Online Social Networking")	984

S53	S51 OR S52	27,039

S54	S50 NOT S53	19,612

S55	S17 OR S18 OR S19 OR S20 OR S21 OR S22 OR S23 OR S24 OR S25 OR S26 OR S27 OR S28 OR S29 OR S30 OR S31 OR S32 OR S33 OR S34 OR S35 OR S36 OR S37 OR S38 OR S39 OR S40 OR S41 OR S42 OR S43 OR S44 OR S45 OR S46 OR S54	337,287

S56	S7 AND S14 AND S55	1,961

S57	(MH "Animals")	94,114

S58	(MH "Human")	2,885,109

S59	S57 NOT (S57 AND S58)	85,610

S60	S56 NOT S59	1,959

S61	S56 NOT S59 - Limiters - English language	1,912

S62	(MH "Literature Review+")	165,106

S63	(MH "Clinical Trials+")	362,101

S64	(MH "Qualitative Studies+")	206,688

S65	S62 OR S63 OR S64	714,320

S66	S61 NOT S65	1,637

S67	EM 20240130-20250423	287,397

S68	S66 AND S67	114

**Source: APA PsycINFO**

Interface: EBSCOhost

Database coverage dates: 1597 to present

Search date: 23 April 2025

Retrieved records: 257

Search strategy:

S1	DE "Attention Deficit Disorder with Hyperactivity"	37,432

S2	TI ( ((attention* or behav*) n2 (defic* or disorder* or dysfunc*)) ) OR AB ( ((attention* or behav*) n2 (defic* or disorder* or dysfunc*)) )	75,617

S3	TI adhd OR AB adhd	34,789

S4	TI "ad/hd" OR AB "ad/hd"	588

S5	TI addh OR AB addh	130

S6	TI adhs OR AB adhs	708

S7	S1 OR S2 OR S3 OR S4 OR S5 OR S6	87,526

S8	AG Childhood	641,171

S9	AG Adolescence	536,612

S10	TI ( (child* or boy* or girl* or toddler* or infant* or pre#school* or schoolchild* or schoolgirl* or schoolboy* or adolescen* or juvenile* or teen* or "young* person*" or "young* people*" or youth* or minor#) ) OR AB ( (child* or boy* or girl* or toddler* or infant* or pre#school* or schoolchild* or schoolgirl* or schoolboy* or adolescen* or juvenile* or teen* or "young* person*" or "young* people*" or youth* or minor#) )	1,191,678

S11	DE "Pediatrics"	36,314

S12	TI ( (pediatric* or paediatric*) ) OR AB ( (pediatric* or paediatric*) )	47,274

S13	S8 OR S9 OR S10 OR S11 OR S12	1,439,650

S14	TI ( (stress* n1 (event* or generat* or dependent or minor or major)) ) OR AB ( (stress* n1 (event* or generat* or dependent or minor or major)) )	16,190

S15	TI oxidative OR AB oxidative	13,582

S16	S14 NOT S15	15,994

S17	TI daily hassle# OR AB daily hassle#	1,286

S18	DE "Social Structure"	6,665

S19	DE "Social Skills"	17,350

S20	DE "Social Support"	71,038

S21	DE "Social Exclusion"	2,595

S22	DE "Social Deprivation"	1,111

S23	DE "Social Isolation"	14,751

S24	DE "Social Capital"	8,400

S25	TI ( (social n (thinning or structure or architecture or group# or experience# or skill# or support or ratings or status or integrat* or relationship# or exclusion or contact# or dysfunction* or depriv* or isolat* or capital)) ) OR AB ( (social n (thinning or structure or architecture or group# or experience# or skill# or support or ratings or status or integrat* or relationship# or exclusion or contact# or dysfunction* or depriv* or isolat* or capital)) )	1,500

S26	TI SNA OR AB SNA	700

S27	TI "personal network" OR AB "personal network"	360

S28	TI (social n1 process*) OR AB (social n1 process*)	18,529

S29	TI socialising OR AB socialising	356

S30	DE "Interpersonal Relationships"	27,703

S31	TI ( ((interpersonal or affiliative or supportive) n relationship#) ) OR AB ( ((interpersonal or affiliative or supportive) n relationship#) )	54

S32	TI ( (shared n (understanding or norms or values or identit*)) ) OR AB ( (shared n (understanding or norms or values or identit*)) )	12

S33	DE "Community Involvement"	7,694

S34	TI ( (community n (participation or inclusion or involve*)) ) OR AB ( (community n (participation or inclusion or involve*)) )	92

S35	TI "Involvement in a life situation" OR AB "Involvement in a life situation"	14

S36	TI participation OR AB participation	120,929

S37	DE "Close Relationships"	1,304

S38	TI ( ((formal or intimate or close) n relationships) ) OR AB ( ((formal or intimate or close) n relationships) )	97

S39	DE "Friendship"	11,709

S40	TI friend* OR AB friend*	83,834

S41	TI ( ((rejected or excluded or disliked) n children) ) OR AB ( ((rejected or excluded or disliked) n children) )	19

S42	DE "Peer Relations"	19,507

S43	DE "Interpersonal Interaction"	34,271

S44	TI ( ((peer# or inter#personal) n1 (accept* or problem# or conflict or likability or liked or status or interact* or function* or relationship# or interaction# or reject*)) ) OR AB ( ((peer# or inter#personal) n1 (accept* or problem# or conflict or likability or liked or status or interact* or function* or relationship# or interaction# or reject*)) )	45,509

S45	DE "Relationship Quality"	8,463

S46	TI ( ((number# or quality or quantity) n1 relationship#) ) OR AB ( ((number# or quality or quantity) n1 relationship#) )	12,146

S47	DE "Bullying"	12,903

S48	DE "Victimization"	28,283

S49	TI ( (victim* or bullied or bully*) ) OR AB ( (victim* or bullied or bully*) )	84,655

S50	TI marginali?ed OR AB marginali?ed	15,507

S51	DE "Loneliness"	9,562

S52	TI lonel* OR AB lonel*	17,554

S53	DE "Popularity"	2,185

S54	TI ( (popular* or unpopular*) ) OR AB ( (popular* or unpopular*) )	65,190

S55	DE "Social Networks"	15,909

S56	DE "Social Network Analysis"	2,068

S57	TI "social network" OR AB "social network"	16,006

S58	S55 OR S56 OR S57	25,670

S59	DE "Social Media"	22,108

S60	DE "Online Social Networks"	10,818

S61	S59 OR S60	29,314

S62	S58 NOT S61	22,716

S63	S16 OR S17 OR S18 OR S19 OR S20 OR S21 OR S22 OR S23 OR S24 OR S25 OR S26 OR S27 OR S28 OR S29 OR S30 OR S31 OR S32 OR S33 OR S34 OR S35 OR S36 OR S37 OR S38 OR S39 OR S40 OR S41 OR S42 OR S43 OR S44 OR S45 OR S46 OR S47 OR S48 OR S49 OR S50 OR S51 OR S52 OR S53 OR S54 OR S62	607,950

S64	S7 AND S13 AND S63	4,944

S65	DE "Animals"	269,782

S66	S64 NOT S65	4,906

S67	DE "Literature Review" OR DE "Systematic Review"	23,953

S68	DE "Clinical Trials" OR DE "Randomized Controlled Trials" OR DE "Randomized Clinical Trials"	14,222

S69	DE "Qualitative Methods" OR DE "Focus Group" OR DE "Grounded Theory" OR DE "Interpretative Phenomenological Analysis" OR DE "Narrative Analysis" OR DE "Semi-Structured Interview" OR DE "Thematic Analysis" OR DE "Focus Group Interview"	23,522

S70	S67 OR S68 OR S69	61,446

S71	S66 NOT S70	4,866

S72	S66 NOT S70 - Limiters - English language	4,568

S73	RD 20240130-20250423	244,059

S74	S72 AND S73	257

### Appendix 2

Risk of bias scoring

Criteria for the scoring of each item of the JBI critical appraisal checklist for cohort studies (<https://doi.org/10.46658/JBIMES-24-01>) were as follows:

- Score = 1 for low risk of bias or when N/A (e.g., if there was no attrition, item #10 would be N/A as it would not be required to deploy strategies to address incomplete follow-up, but this would lower the total score of the appraisal, hence we scored N/A as 1);
- Score = -1 for high risk of bias, i.e., the item was not addressed adequately;
- Score = 0 if evidence of the item being addressed is not clearly outlined.

Item #6 of the JBI checklist was excluded from the calculation, hence the “applicable score” was calculated dividing the total score by the 10 items considered.

### Appendix 3

##### JBI quality appraisal table

| **Study** | **1** | **2** | **3** | **4** | **5** | **6** | **7** | **8** | **9** | **10** | **11** | **Score** |
| --- | --- | --- | --- | --- | --- | --- | --- | --- | --- | --- | --- | --- |
| Murray-Close D. *et al*., 2010^37^ | ✗ | ? | ✓ | ✓ | ✓ |  | ✓ | ✓ | ✓ | ✓ | ✓ | 7 |
| Owens E. B. *et al.*, 2009^38^ | ✓ | ✓ | ✓ | ✓ | ✓ |  | ✓ | ✓ | ✓ | NA | ✓ | 10 |
| Mikami A. Y. *et al.*, 2015^39^ | ✗ | ✓ | ✓ | ✓ | ✓ |  | ✓ | ✓ | ✓ | ✓ | ✓ | 9 |
| Lee S. S. *et al.*, 2008^40^ | ✓ | ✗ | ✓ | ✓ | ✓ |  | ✓ | ✓ | ? | ✓ | ✓ | 7 |
| Efron D. *et al.*, 2020^41^ | ✓ | ✓ | ✓ | ✓ | ✓ |  | ✓ | ✓ | ✓ | ✗ | ✓ | 9 |
| Biederman J., 1996^42^ | ✓ | ✓ | ✓ | ✓ | ✓ |  | ✓ | ✓ | ✓ | NA | ✓ | 10 |
| Reinke A. L. *et al.*, 2023^43^ | ✓ | ✓ | ✓ | ✓ | ✓ |  | ✓ | ✓ | ✗ | ✓ | ✓ | 9 |
| Efron D. *et al.*, 2021^44^ | ✓ | ✓ | ✓ | ✓ | ✓ |  | ✓ | ✓ | ✓ | ✓ | ✓ | 10 |
| Fogelman N. D. *et al.*, 2018^45^ | ✓ | ✓ | ✓ | ✓ | ✓ |  | ✓ | ✗ | ✓ | ? | ✓ | 7 |
| Hennig T. *et al.*, 2017^46^ | ✓ | ✓ | ✓ | ✓ | ✓ |  | ✓ | ✓ | ✗ | ✗ | ✓ | 6 |
| Lin C.-J. *et al.*, 2024^47^ | ✓ | ✓ | ✓ | ✓ | ✓ |  | ✓ | ✓ | NA | NA | ✓ | 10 |

Legend: ✓ = Yes, ✗ = No, ? = Unclear, NA = Not Applicable.

JBI critical appraisal checklist for cohort studies - complete questions:

1. Were the two groups similar and recruited from the same population?
2. Were the exposures measured similarly to assign people to both exposed and unexposed groups?
3. Was the exposure measured in a valid and reliable way?
4. Were confounding factors identified?
5. Were strategies to deal with confounding factors stated?
6. Were the groups/participants free of the outcome at the start of the study (or at the moment of exposure)?
7. Were the outcomes measured in a valid and reliable way?
8. Was the follow up time reported and sufficient to be long enough for outcomes to occur?
9. Was follow up complete, and if not, were the reasons to loss to follow up described and explored?
10. Were strategies to address incomplete follow up utilized?
11. Was appropriate statistical analysis used?
